# Antibiotic resistance genes, antibiotic residues, and microplastics in influent and effluent wastewater from treatment plants in Norway, Iceland, and Finland

**DOI:** 10.1101/2025.02.21.25322651

**Authors:** Ananda Tiwari, Adrián Jaén-Gil, Anastasia Karavaeva, Alessio Gomiero, Ásta Margrét Ásmundsdóttir, Maria João Silva, Elisa Salmivirta, Thanh Tam Tran, Anniina Sarekoski, Jeremy Cook, Rolf Lood, Tarja Pitkänen, Adriana Krolicka

## Abstract

Monitoring antimicrobial resistance genes (ARGs) in wastewater influent (pre-treatment) and effluent (post-treatment) reveals their circulation in communities via wastewater surveillance (WS), possible amplification during treatment, and potential public health risks from gene releases into surface water. This study used Oxford Nanopore (ONP) metagenomic sequencing and qPCR to track ARGs in wastewater treatment plants (WWTPs) influents and effluents in Mekjarvik (Norway), Reykjavik (Iceland), and Mariehamn (Åland-Finland). High-Performance Liquid Chromatography (HPLC) monitored antibiotic residues and Micro-Fourier Transform Infrared Spectroscopy (µ-FTIR) used for microplastics (MPs) in Mekjarvik and Reykjavik. Metagenomic analysis identified 193 unique ARGs, with the highest average (±SD) in Reykjavik (66.3 ± 4.1), followed by Mekjarvik (61.3 ± 14.1) and Mariehamn (18.0 ± 2.2). While treatment generally reduced ARGs, ONP and qPCR detected amplification of some carbapenemase and ESBL genes. ONP sequencing linked many ARGs to plasmids, co-occurring with metal stress genes. The most prevalent plasmids—*Col440I*, *IncQ2*, and *ColRNAI*—were found across all WWTPs. Mercury-related genes dominated metal stress genes (64.9%), followed by multimetal (23.7%) and copper (6.4%) stress genes. Among 45 antibiotics screened in Mekjarvik and Reykjavik, only sulfamethoxazole and sulfapyridine were consistently quantified, while azithromycin, ciprofloxacin, and ofloxacin were often below detection limits. MPs were highest in Reykjavik influent (8200 MPs/m³) and Mekjarvik influent (5900 MPs/m³). Treatment effectively reduced larger MPs but was less effective against smaller particles. Polyethylene (∼60%) was the most dominant MP type, except in Mekjarvik influent, where polypropylene (∼50%) prevailed. This study reveals distinct ARG and antibiotic residue patterns in wastewater. While treatment significantly reduced ARGs, antibiotic residues, and larger MPs, it did not eliminate them, posing risks for environmental pollution. ARGs related to carbapenemase and ESBL persisted, and fine MPs increased post-treatment. These findings underscore the need to monitor both influent and effluent to have information about removal efficiencies and needs to protect downstream water environments. The detection of diverse ARGs, plasmids, and genes of critical pathogens like *Acinetobacter baumannii*, *Pseudomonas aeruginosa*, *Escherichia coli*, and *Salmonella* spp. in wastewater effluent presents a significant environmental challenge and emerging pollutant for recipient waters.

**Highlights:**

- Influent and effluent wastewater samples from Norway, Iceland, and Finland were analyzed.
- Resistance genes were analyzed via high-throughput qPCR and Oxford Nanopore (ONP) metagenomics.
- Dominant ARG groups in Mekjarvik and Mariehamn were tetracycline and macrolide but Quinolone, and macrolide in Reykjavik.
- Only sulfamethoxazole and sulfapyridine were consistently detected in Mekjarvik and Reykjavik, out of 45 screened antibiotics.
- Treatment effectively removed larger MPs but was less effective against smaller ones.

## 1. Introduction

Antimicrobial resistance (AMR) poses a serious global public health threat, resulting in millions of cases of illness and death annually^1^. Wastewater surveillance (WS), i.e. monitoring wastewater influents, is an emerging tool for assessing AMR at a population level ^2–5^. Municipal wastewater can be an ideal material for WS as it receives a wide range of pathogens from households and healthcare settings that end up through various excretion routes (feces, urine, nasal secretions, saliva, and skin lesions) from a wide range of individuals (symptomatic, asymptomatic, pre-symptomatic, and post-symptomatic)^4,6^. WS offers nearly real-time evidence of infectious agents or their genetic material in sewage systems, as soon as shedding the pathogens and markers in sewage system, even before symptoms appear and seeking clinical testing^6,7^.

Expanding and improving sewage collection and treatment through wastewater treatment plants (WWTPs) is an effective way to curb the spread of resistant microorganisms via environmental pathways ^8^. The ecological interaction of microbial communities during the treatment process may affect the prevalence of antimicrobial-resistant bacteria (ARB) and antimicrobial-resistance genes (ARGs) ^8,9^. WWTPs receive various antimicrobial agents, including traces of partially metabolized and discarded antibiotics, heavy metals, and detergents ^8^. These compounds create selective ecological pressure on bacterial communities for emerging resistant communities. Biological treatment processes, specifically, activated sludge, enhance microbial growth and create an ideal environment for horizontal gene transfer (HGT) of ARGs via mobile genetic elements (MGEs) like plasmids and transposons, making wastewater a hotspot for the proliferation of ARGs ^8,10^. Consequently, WWTPs serve as critical sinks and reservoirs, as well as major sources of antimicrobial residues, ARGs, and AMR in recipient surface waters ^10^.

Microplastics (MP) are emerging environmental pollutants from sources like personal care products, synthetic fibers, and degrading plastic debris^11,12^. They threaten aquatic ecosystems and human and animal health by spreading through contaminated food and water^11,13^. The relationship between MP and ARGs in wastewater treatment is complex but significant^13^. Studies suggest MPs create a “plastisphere” for bacterial communities and resistant pathogens, fostering biofilms^13,14^. Their presence in WWTPs can influence microbial communities, potentially affecting ARG removal^14^. The EU’s Urban Wastewater Treatment Directive (UWWTD) encourages advanced treatment technologies to reduce MPs and microbial contaminants, including ARGs, improving effluent quality^15^. Water connects humans, animals, and the environment, making it an important link in the One Health concept^16^. The release of clinically relevant ARB and ARGs into the environment poses significant health risks to humans and animals, underscoring the need for effective wastewater treatment and reducing pollutant loads into recipient water ^4,8,17^.

The SOS-*E. coli* assay is a bioanalytical method for quickly evaluating WWTP efficiency in removing micropollutants (e.g., antibiotics) that stress bacteria and contribute developoing antimicrobial resistance^11,12^. It assesses the genotoxicity and stress response of *Escherichia coli* strains exposed to environmental toxicants like antibiotics^11^. The assay detects the activation of the SOS response, a cellular mechanism triggered by DNA damage or stress, leading to the induction of repair proteins and stress-related genes^11,12^.

Molecular methods like PCR/qPCR offer high sensitivity for detecting both culturable and hard-to-culture strains using specific primers and probes for pathogens and ARGs^8,18^.

Traditional qPCR monitors only a few targets per run, making large-scale monitoring time-consuming and labor-intensive. In contrast, high-throughput qPCR (HT-qPCR) can analyze dozens of targets in a single run, has been applied in hospital wastewater^19^ and municipal wastewater^20,21^. Also, classical qPCR requires fixed labs and a stable power supply, making on-site testing impractical and delaying responses due to sample transport. The Biomeme device offers a portable, battery-powered qPCR platform for on-site testing at WWTPs or remote locations, delivering results within hours and reducing errors and transport costs^22,23^. It also integrates with cloud-based platforms for real-time data sharing, enabling instant access for multiple stakeholders^22,23^. Metagenomics, on the other hand, is a robust screening tool that does not require prior knowledge of primers and probes, making it useful for tracking and screening emerging ARGs ^2,8^. Recently, Oxford Nanopore (ONP) sequencing has gained popularity due to its versatility in genome assembly, detecting long-length transcripts, and identifying mutations and genomic changes ^24^. Its popularity stems also from portability, quick turnaround time, potential on-site testing, low capital cost, and minimal infrastructure requirements ^24,25^. ONP shows promise for real-time pathogen detection and surveillance, potentially vital for pandemic response. The compact MinION device allows for onsite high-throughput sequencing directly in field conditions.

This study monitored ARGs via qPCR and ONP in the influent and effluent of three WWTPs: Mekjarvik (Norway), Klettagarðar Reykjavik (Iceland), and Mariehamn Åland (Finland). Antibiotic residues and microplastics were analyzed only in Mekjarvik and Reykjavik. The study tested the following hypotheses: (a) Influent resistome composition reflects regional ARG patterns; (b) ARG removal efficiencies vary across resistance classes and WWTPs; (c) qPCR- and metagenomics-based ARG monitoring methods show concordance; (d) ARG prevalence correlates with antibiotic traces in wastewater. We anticipate this study will support the EU’s Urban Wastewater Treatment Directive (UWWTD) ^26^, and the development of a unified ARG WS system in Nordic countries ^27^. It may contribute to harmonizing results from ONP metagenomics and qPCR-based methods and understanding the impact of microplastics on ARGs in wastewater.

## 2. Methods

### 2.1 Site description, and sample collection

Wastewater samples were collected from three WWTPs — Mekjarvik (Norway), Klettagarðar Reykjavik (Iceland), and Mariehamn (Åland, Finland) — representing diverse Nordic regions. Mekjarvik WWTP (Nord-Jæren) is one of the largest treatment plants in Norway on the southwestern coast, serving about 300,000 people from Randaberg, Stavanger, Sola, Sandnes, and Gjesdal. It is a full-scale treatment plant consisting of primary and secondary treatment units, effectively removing phosphorus based on chemical precipitation. The plant has been operated since 1991, fully in compliance with the UWWTD 1991 of the European Union ^28^.

The Klettagarður WWTP in Reykjavík, Iceland, is the country’s largest plant, operating since 2002 ^29^. It serves ∼ 160,000 residents and several key industrial areas. It relies solely on primary treatment, using 3 mm filter before discharge ^29^. Such filtered effluent, diluted with drainage water and discharged 5,500 meters offshore at a depth of about 30 meters. The final 1,000 meters of the discharge pipe are perforated with 7 cm holes to enhance mixing and dilution. Under typical dry conditions, the plant processes an average flow of 1,500 L/s, which can double during wet weather ^29^.

Mariehamn WWTP, located in Åland, Finland—serves around 21,000 people from Mariehamn, Jomala, Lemland, Hammarland, Finström, Saltvik, and Sund ^30^. The plant has advanced facilities, with both primary and secondary treatment units, compliant with the UWWTD 1991 of the EU ^28^. More details on the Mekjarvik ^31^, Reykjavík ^29^, and Mariehamn ^30^ WWTPs have been published earlier.

Monthly 24-hour composite influent and effluent samples were collected between June 2023 and February 2024. Sample collection and processing followed standardized protocols to ensure preservation and stability during shipment ^21^. Samples for molecular analysis were collected in sterile polypropylene containers, Reykjavik samples were stored below -75°C and shipped to Norwegian Research Centre’s (NORCE) Chemical-microbiology lab in Stavanger, Norway. The Mekjarvik WWTP is located about 200 meters from NORCE’s microbiology lab in Stavanger, Norway. Mariehamn samples were sent to the Finnish Institute for Health and Welfare’s (THL) microbiology lab in Kuopio, Finland ^30^. Samples were transported in a cool box to the laboratory, maintaining a low temperature, and analyzed within 24 hours.

### 2.2 Bacterial community analysis and ARGs monitoring

#### 2.2.1 Wastewater concentration and DNA extraction

Triplicate aliquots of each sample, influent (35 mL), and effluent samples (240 mL) were filtered using a Sterivex-GP Pressure Filter Unit (0.22 µm pore size, polyethersulfone membrane, Millipore) for bacterial biomass concentration. Aseptically, filters were torn into ∼1 cm² pieces with sterile forceps and placed in extraction tubes with 1.5 mL of 3M guanidine thiocyanate lysis solution (pH 8.9) and incubated at 85 °C for 10 minutes in a ThermomixerC at 500 rpm. The lysate was filtered through a 0.2 µm syringe filter (25 mm Cellulose Acetate, VWR cat no. 514-1273) into a Falcon tube to minimize foam. The lysate was transferred to the M1 Sample Prep Cartridge Kit (Biomeme #3000536R). DNA purification was performed according to the cartridge manual with minor modifications. To increase the total concentration and purity of DNA (1) 650 µL elution buffer from the last section of the cartridge was removed, DNA was eluted using approximately 150 µL elution buffer (2) Prior air dying procedure after wash step the syringe and column were shaken vigorously to remove any remaining alcohol used during the washing step.

#### 2.2.2 Oxford Nanopore metagenomics

Samples collected over three sampling events from Mekjarvik (May–July), Reykjavik (May– July), and Mariehamn (May, August, September) were used for ONP metagenomic sequencing. Composite samples were prepared by pooling triplicates with equal DNA mass. Multiple displacement amplification (MDA) to enrich the whole metagenome to ensure a sufficient amount of total DNA, was performed using high fidelity EquiPhi29 DNA Polymerase and 5′-phosphorylated random hexamers (ThermoFisher# A65393). The reaction was performed according to the manufacture instruction. The DNA library was constructed using the Rapid Barcoding Kit (SQK-RBK114-96) and loaded onto an R10.4.1 flow cell for sequencing on a MinION device. Raw sequencing data were processed into nucleotide sequences and filtered for low-quality reads using MinKNOW software in high-accuracy mode. The NanoFilt tool was used for preprocessing, including the removal of reads shorter than 1000 base pairs, reads with an average quality score lower than 9, and the removal of first 50 bases from each read, to ensure data quality. Human reads were identified and discarded using a combination of the Minimap2 and Kraken2 tools ^32,33^. Reads mapped to the human genome reference GRCh38 by Minimap2 were discarded, followed by running the extract_kraken_reads.py script on the initial Kraken2 results to filter out the remaining reads belonging to taxid 9443 (human) ^32,33^. Taxonomic classification of the metagenome was performed using Kraken2 with the PlusPF-8 database ^33,34^. Microbial abundances were standardized as a percentage of total genera per sample.

The ABRicate tool, integrated into the Epi2me pipeline, along with the Resfinder database was employed to identify ARGs ^35^. Results were presented with and without normalization using gigabase pair sequences. As the “read count” provided by the Epi2me Labs output acts as the normalization factor, the read counts per ARG was directly used as abundance values for downstream analyses, eliminating the need for additional normalization methods, such as ARG copies per gene, ARG copies per 16S rRNA gene, or relative abundances. The ABRicate tool together with the PlasmidFinder database, was used to identify plasmids ^36^. Through PlasmidFinder, we identified instances where the same read IDs matched ARGs detected by Resfinder. Such co-detection strongly indicates that the ARGs are likely plasmid-borne. For additional analyses, custom databases were incorporated into ABRicate. Metal resistance genes were identified using the newer Megares v3.0 database, while MGEs were detected using databases such as ISfinder ^37^. Metagenomes are publicly available through the National Center for Biotechnology Information’s BioProject PRJNAxxx.

#### 2.2.3 Biomeme qPCR analysis

ARGs from the Mekjarvik (Norway) and Reykjavik (Iceland) WWTPs were monitored using the Franklin™ Real-Time PCR Thermocycler (Biomeme) platform. Triplicate samples were collected monthly in Reykjavik from May 2023 to April 2024, except for November 2023. In Mekjarvik WWTP, monthly triplicate samples were collected from July 2023 to January 2024 for Biomeme qPCR analysis. Standard curves for the analyzed genes (Table 1) were established using gBlocks Gene Fragments (Integrated DNA Technologies, IDT, Coralville, IA, USA). All reactions were conducted using the standard Biomeme LyoDNA program, with an annealing temperature of 60 °C, a primer concentration of 500 nM, and 5 µL of DNA. Samples were analyzed using LyoGreen™ 2.0 Master Mix (Biomeme). Positive controls constituted reactions performed using Mastemix LyoDNA™ 2.0 + IPC Master Mix to identify any potential inhibition effects.

**Table 1.** Summary of Microbial reads and Diversity Across Wastewater Treatment Plants.

| WWTP name | Sample name | Total number of microbial reads | Bacterial reads | Total number of species | Total number of Genera | Total number of class |
| --- | --- | --- | --- | --- | --- | --- |
| Mekjarvik (Norway) | Influent_May | 182,790 (12.5%) | 182,324 (12.6%) | 3598/3398 | 1224/1086 | 107/66 |
|  | Influent_June | 104,447 (7.1%) | 104,203 (7.2%) | 2488/2381 | 917/836 | 91/56 |
|  | Influent_July | 101,624 (6.9%) | 101,378 (7.0%) | 2629/2506 | 955/865 | 90/58 |
|  | **Influent Total** | **388,861 (26.5%)** | **387,905 (26.8%)** | **5094/4812** | **1567/1390** | **124/77** |
|  | Effluent_May | 152,373 (10.4%) | 144481 (10.0%) | 4787/4510 | 1469/1302 | 112/68 |
|  | Effluent_June | 68,664 (4.7%) | 64292 (4.4%) | 2940/2791 | 1097/983 | 101/60 |
|  | Effluent_July | 175,124 (11.9%) | 174752 (12.1%) | 3231/3080 | 1084/982 | 91/55 |
|  | **Effluent Total** | **396,161 (27.0%)** | **383525 (26.5%)** | **6126/5749** | **1744/1535** | **123/75** |
| Reykjavik (Iceland) | Influent_May | 89,744 (6.1%) | 89,553 (6.2%) | 2539/2449 | 894/818 | 75/52 |
|  | Influent_June | 76,892 (5.2%) | 76,738 (5.3%) | 2445/2357 | 865/794 | 83/52 |
|  | Influent_July | 138,343 (9.4%) | 138,131 (9.5%) | 2892/2796 | 991/913 | 86/57 |
|  | **Influent Total** | **304,979 (20.8%)** | **304,422 (21.0%)** | **4412/4216** | **1382/1245** | **104/67** |
|  | Effluent_May | 123,614 (8.4%) | 123,409 (8.5%) | 2982/2871 | 1029/937 | 87/56 |
|  | Effluent_June | 54,892 (3.7%) | 54,752 (3.8%) | 2367/2296 | 820/757 | 80/53 |
|  | Effluent_July | 48,492 (3.3%) | 48,373 (3.3%) | 2030/1960 | 718/660 | 76/50 |
|  | **Effluent Total** | **226998 (15.5%)** | **226,535 (15.7%)** | **4219/4037** | **1319/1188** | **104/67** |
| Mariahamn (Åland-Finland) | Influent _ May | 53,553 (3.7%) | 53,124 (3.7%) | 3062/2925 | 1127/1029 | 102/68 |
|  | Influent _ Aug | 55,452 (3.8%) | 54,958 (3.8) | 3300/3136 | 1198/1097 | 101/70 |
|  | Influent _ Sept | 20,612 (1.4%) | 20,213 (1.4%) | 2258/2129 | 968/880 | 101/64 |
|  | **Influent Total** | **129,617 (8.8%)** | **129,617 (8.9%)** | **5193/2050** | **1652/1479** | **132/86** |
|  | Effluent_May | 7,877 (0.5%) | 7,348 (0.5%) | 2195/2050 | 924/812 | 96/65 |
|  | Effluent_Aug | 6,739 (0.5%) | 6,411 (0.4%) | 2344/2201 | 989/885 | 103/69 |
|  | Effluent_Sept | 5,063 (0.3%) | 4,854 (0.3%) | 1490/1401 | 716/649 | 81/54 |
|  | **Effluent Total** | **19,679 (1.3%)** | **18,613 (1.4%)** | **4259/3990** | **1471/1294** | **120/79** |
|  | **Granda**<br>**Total** | **1,466,295** | **1,449,294** | **8434/7709** | **2195/1813** | **152/93** |

**Note:** Percentage in Total number of microbial read and Bacterial reads columns are calculated out of Grand Total reads in 18 samples

#### 2.2.4 High-throughput qPCR

The prevalence of ARGs in the Mariehamn (Åland, Finland) sample was investigated with a SmartChip Real-Time PCR system (Takara Bio, Mountain View, CA, USA), by Resistomap Oy (Helsinki, Finland) (https://www.resistomap.com/). Nucleic acid concentration from each sample was diluted to 10 ng/µl with nuclease-free water and 300 ng of each sample was shipped in cool boxes with ice within 24-hours. The samples were fitted on 48 sample-based chips, together with a negative control, one NTC, and one positive control ^19,21^. Reagents and qPCR cycling conditions described in Wang et al. (2014) ^21^. As described in Sarekoski et al. to minimize the possibility of false-positive or false-negative results, each sample was run in three replicates, and amplification in at least two out of the three replicates was considered positive^21^. The cycle threshold (CT) was set to 27 and the mean value of the replicates was used for analysis.

Table S2 lists the assays used in the SmartChip qPCR system to analyze ARGs and taxonomic markers in wastewater samples from Mariehamn, Åland, Finland. The analysis screened 21 clinically relevant ARGs, including those conferring resistance to wide range of beta-lactams, carbapenemases, and *ampC* (*bla*_CTX-M_*, bla*_CMY_*, bla*_MOX_*/bla*_CMY_*, ampC/bla*_DHA_*, bla*_ACT_*, bla*_OXA-10_*, bla*_OXA23_*, bla*_OXA48_*, bla*_OXA-58_*, bla*_SME_*, bla*_VEB_*, bla*_VIM_*, bla*_GES_*, bla*_KPC_), vancomycin (*vanA, vanB*), and tetracycline (tetO_1). Taxonomic markers (crAssphage assay *crAss56, A. baumannii, K. pneumoniae, Staphylococci* spp.) were also targeted. The *crAss56* assay served as a positive control, and the relative abundance of ARGs was determined by normalizing cycle threshold (CT) values against the 16S rRNA gene CT value (ΔCT = CT targeted gene – CT 16S rRNA gene) ^19,21,38^.

### 2.3 Antibiotic residues monitoring

#### 2.3.1 Sample preparation and preconcentration

Duplicate samples of 100 mL influent wastewater and 150 mL effluent wastewater were initially spiked with a mixture of deuterated standards, including azithromycin-d^3^, ofloxacin-d_8_, and sulfamethoxazole-d^4^ (Toronto Research Chemicals, Canada), at a final concentration of 300 ng/L. The spiked samples were then processed using the solid-phase extraction (SPE) methodology as previously described ^39^. Briefly, the samples were first filtered through 1 µm glass fiber filters and subsequently through 0.45 µm PVDF membrane filters (Millipore, Billerica, MA, USA). The SPE cartridges Oasis HLB (200 mg, 6 mL) from Waters Corporation (Milford, MA, USA) were conditioned with 6 mL of methanol, followed by 5 mL of pure water. The filtered samples were then loaded onto the cartridges, rinsed with 6 mL of pure water, and dried for 30 min to remove residual water. Elution was performed with 7 mL of methanol. The eluate was dried under nitrogen and reconstituted in 1 mL of a methanol/water solution (10:90, v/v).

#### 2.3.2 Suspect screening analysis

A total of 45 antibiotics were screened (Table S3). Antibiotic analysis was conducted using an adapted suspect screening method ^39^, with a liquid chromatography system coupled to an LC-Orbitrap Exploris 240 high-resolution mass spectrometer (Thermo Fisher Scientific). Chromatographic separation used a ZORBAX Eclipse Plus C18 (150 mm × 2.1 mm, 3.5 µm; Agilent Technologies) at 0.350 mL/min for a 20-minute run. The mobile phase selected were: (A) water: methanol (98:2) with ammonium formate 5 mM and acid formic 0.01% and (B) methanol: water (98:2) with ammonium formate 5 mM and acid formic 0.01%. The solvent gradient started with an initial mobile phase composition of 95% A and held for 1 min, followed by a decrease in composition A to 5% within 11 min and held for 2 min, and finally up to 95% in 1 min and held for 5 min.

The Orbitrap Exploris 240 was equipped with a heated electrospray ionization source (HESI-II), and the analyses were performed in a positive mode^40^. Samples were acquired in Data Dependent Acquisition (DDA) mode through full-scan at a mass-to-charge (*m/z*) from 100 to 1000 and a resolving power of 60.000 FWHM, followed by a second event from *m/z* 50 to 1000 range at 30.000 FWHM. The most intense ions (Top 5) from the data scan were selected from an inclusion list of tentative antibiotic exact masses (Supplementary Material X), and fragmented using high-collision dissociation (HCD) at a collision energy of 40 eV and an isolation width of 2 Da. The entire system was controlled via Thermo Scientific Xcalibur (Version 4.4.16.14) software.

#### 2.3.3 Computational data treatment and semi-quantification

The Compound Discoverer 3.3 SP2 software (Thermo Fisher Scientific) was used for the identification of the suspected antibiotic residues in recorded files. Automatic data processing started with selecting the MS spectra from *m/z* 100 to 1000 and from 1 to 14 min with a peak intensity threshold at 1.5 signal-to-noise ratios (S/N). The antibiotic residues present in the inclusion list (Table S3) were added as expected compounds to be detected in collected data using a mass tolerance error of ± 5 ppm, a minimum peak intensity of 1000 counts, and a 1.5 chromatographic S/N threshold. Grouping expected compounds was performed at a retention time (Rt) tolerance of 0.3 min and a mass tolerance of ± 5 ppm for merging features. The fragment spectra were calculated using FISh scoring at an S/N threshold of 3 and a mass tolerance error of ± 5 ppm. To ensure the reliability of identification, the generated data was manually reviewed and compared with the internal standards of each antibiotic (if available). Semi-quantification was performed using the most similar internal standard in terms of chemical structure and chromatographic retention time as described earlier ^41^.

### 2.4 Monitoring of microplastics

#### 2.4.1 Samples collection

For microplastic (MP) monitoring, wastewater samples from both influent and effluent in Mekjarvik (Norway) and Reykjavik (Iceland) were pumped from the water stream into a custom-designed sampling device. This device consists of a cascade of removable laser-cut stainless-steel filters with mesh sizes of 300 µm and 10 µm, coupled with a peristaltic Oberdorfer bronze rotary head pump. Each sampling event lasted from 1/2 to 2 hours, depending on the amount of suspended material in the wastewater, with a maximum flow rate of 8–10 L/min. The total volume of filtered water was recorded. After each session, both filters were removed from the gaskets, transferred to a pre-cleaned large glass Petri jar, and stored in dark, cold conditions (+4°C) in preparation for sample analysis.

#### 2.4.2 Samples preparation and analysis

The material trapped on the filter was dried in an oven at 60°C for 72 hours. To remove natural organic and inorganic materials before the characterization and quantification of plastic fragments—without damaging the polymer structure—samples underwent enzymatic digestion followed by iron-catalyzed oxidation ^42^. All enzymes (Sigma Aldrich, Germany) were of technical grade and filtered (0.2 µm) before use. Purification began with the addition of 50 mL of sodium dodecyl sulfate (SDS, 5% w/v) per gram of dry weight (DW) sample. Samples were placed in a pre-cleaned 2L crystallization dish and incubated at 50°C for 6 hours. SDS-incubated samples were vacuum-filtered on a pre-muffled 47 mm, 10 µm mesh filter. The digestates were vacuumed off, while the remaining material trapped on the stainless-steel filter was rinsed with 50 mL of GF/A (1.2 µm) filtered ultrapure water and the filter placed on a pre-cleaned 50 mL beaker. Next, 10 mL of a Vicozyme and Cellulase mixture (1:10, v) in acetate buffer (0.1 M; pH 4.8) was added ^42^. The sample was placed in an ultrasonic bath for 5 minutes to facilitate the detachment of the particles from the filter surface and the filter was removed afterwards. The resuspended particles were incubated for 36 hours at 50°C. The digestates were vacuumed off using the same stainless-steel filter, and the digested residual material was rinsed again with 50 mL of ultrapure water. The stainless-steel filter was placed in beaker with 50 mL of protease per gram of DW (Sigma, Germany) in PBS (1:10 at pH 7.4) and submitted to ultrasonic bath for 5 minutes to facilitate the detachment of particles from the stainless-steel membrane and the filter removed from the beaker. The obtained suspension was digested for 48 hours at 30°C, followed by vacuuming and rinsing with ultrapure water. Finally, the residual filter-trapped organic material was once again resuspended using ultrasonication and oxidized using an iron (II)-catalyzed hydrogen peroxide (H_2_O_2_) reaction (Fenton). H_2_O_2_ was added to a final concentration of 250 g/L, along with iron (II), 2.5 g/L ^43^. The filter in the beaker was kept in a water bath to maintain a temperature between 30 and 40°C. The oxidized material was vacuumed off, and the residual iron catalyzer was washed from the stainless-steel filter surface by flushing with 100 mL of 0.5 M HCl solution, followed by 50 mL of Milli-Q water. The obtained sample was transferred to a pre-cleaned separator funnel, and microplastic particles were density-separated from the residual material using a ZnCl_2_ (ρ = 1.75 g/cm³) solution for 10 days. Floating particles were filtered and resuspended in a 5 mL ethanol/water (50:50) solution, and three 0.5 mL aliquots were deposited on a 13 mm Ø Anodisk filter for µ-FTIR scanning by Nicolet iN 10 MX (Thermofisher, Germany). The entire surface of each Anodisk filter was scanned. The scan images and the associated IF spectra were recorded and compared to the polymer database spectra using “SiMPLE - Systematic Identification of MicroPlastics in the Environment” software (Primpke et al., 2020). Each MP was detected and all research scores in % were noted but only polymers matching with a threshold greater than 85% to reference spectra present in SiMPLE database were considered. Each identified particle will be characterized by the software for its major, minor and Ferret dimension as well as the estimated volume and total weight. Three technical replicates were run for each sample; measurements were performed in reflection mode.

### 2.5 SOS-*E. coli* analyses

This study used SOS *E. coli* assay to assess the genotoxicity and stress response of *E coli*—a cellular mechanism triggered by DNA damage or stress, often involving the induction of repair proteins and other stress-related genes. *E. coli* expressing the *recA*-GFP reporter system (Horizon Discovery, PEC3876-202384786) was cultured overnight in Luria-Bertani (LB) medium supplemented with 25 µg/mL kanamycin ^44^. The culture was diluted in LB and grown to an OD600 of ∼0.4, then further diluted to OD600 0.1. Water samples were mixed with the bacteria at a 1:1 (v/v) ratio and incubated at 37°C with shaking at 50 rpm for 1, 2, and 4 hours^44^. Fluorescence was measured using a Victor reader (485/530 nm). All experiments were performed in biological triplicates and technical quadruplicates.

### 2.6 Statistical Analysis

Differences in ARG abundance between groups (spatial variation and influent/effluent) were tested with the Mann-Whitney U test (independent samples) in SPSS. To compare resistome profiles across samples, ARG abundances were standardized as a percentage of total ARGs per sample, and a Bray-Curtis resemblance matrix was generated in Primer-E (version 6.1.13). Non-metric multidimensional scaling (NMDS), Permutational Multivariate Analysis of Variance (PERMANOVA) analyzed beta diversity, assessing differences in microbial communities and antimicrobial resistance genes. NMDS visualized community variation using Bray-Curtis metrics, and PERMANOVA tested statistical significance among three treatment plants, and influent vs. effluent.

We analyzed variations in ARG read counts between influent and effluent samples. ARGs were considered attenuated if their read counts were 2.5 times higher in the influent than in the effluent or if they exceeded three in the influent but were undetected in the effluent. Conversely, ARGs were classified as amplified if their read counts were 2.5 times higher in the effluent than in the influent or if they exceeded three in the effluent but were undetected in the influent. Figures were created with OriginPro.

## 3. Results

### 3.1 Prevalence of Microbial Reads and Diversity via Oxford Nanopore Sequencing

Microbial reads and taxonomic diversity (genera, species, and classes) on each sample in influent and effluent, from three WWTPs are shown in Table 1. A total of 1,466,295 microbial reads were recorded, 98.8% of which were bacterial. Mekjarvik’s influent had 388,861 reads (99.8% bacterial) and effluent 396,161 (96.8% bacterial). Reykjavik’s influent had 304,979 reads and effluent 226,998, both 99.8% bacterial. Mariehamn’s influent had 129,617 reads (99.0% bacterial) and effluent 19,679 (94.6% bacterial). Mekjarvik samples accounted for over half of the total reads, while Mariehamn contributed only about 10% (Table 1).

Microbial communities differed across the three WWTPs (NMDS, PERMANOVA R² = 0.49, p < 0.001). Influent and effluent communities were distinct but showed some overlap (NMDS, PERMANOVA R² = 0.16, p = 0.028) (Figure 1). Across all WWTPs, 8,434 species (91.4% bacterial), 2,195 genera (82.6% bacterial), and 152 classes (61.2% bacterial) were identified. Mekjarvik WWTP had the highest microbial diversity, followed by Mariehamn, with Reykjavik having the lowest (Table 1). In Mekjarvik WWTP, influent contained 5,094 species (94.5% bacterial), 1,567 genera (88.7% bacterial), and 124 classes (62.1% bacterial). Mekjarvik effluent showed slightly higher diversity with 6,126 species (93.8% bacterial), 1,744 genera (88.0% bacterial), while class distribution remained similar (123 total, 61.0% bacterial). In Reykjavik WWTP, influent had 4,412 species (95.6% bacterial), 1,382 genera (90.1% bacterial), and 104 classes (64.4% bacterial), decreasing slightly in effluent to 4,219 species, 1,319 genera, and 104 classes. In Mariehamn WWTP, influent contained 5,193 species (39.5% bacterial), 1,652 genera (89.5% bacterial), and 132 classes (65.2% bacterial). Effluent showed reduced diversity, with 4,259 species, 1,471 genera, and 120 classes.

**Figure 1.**
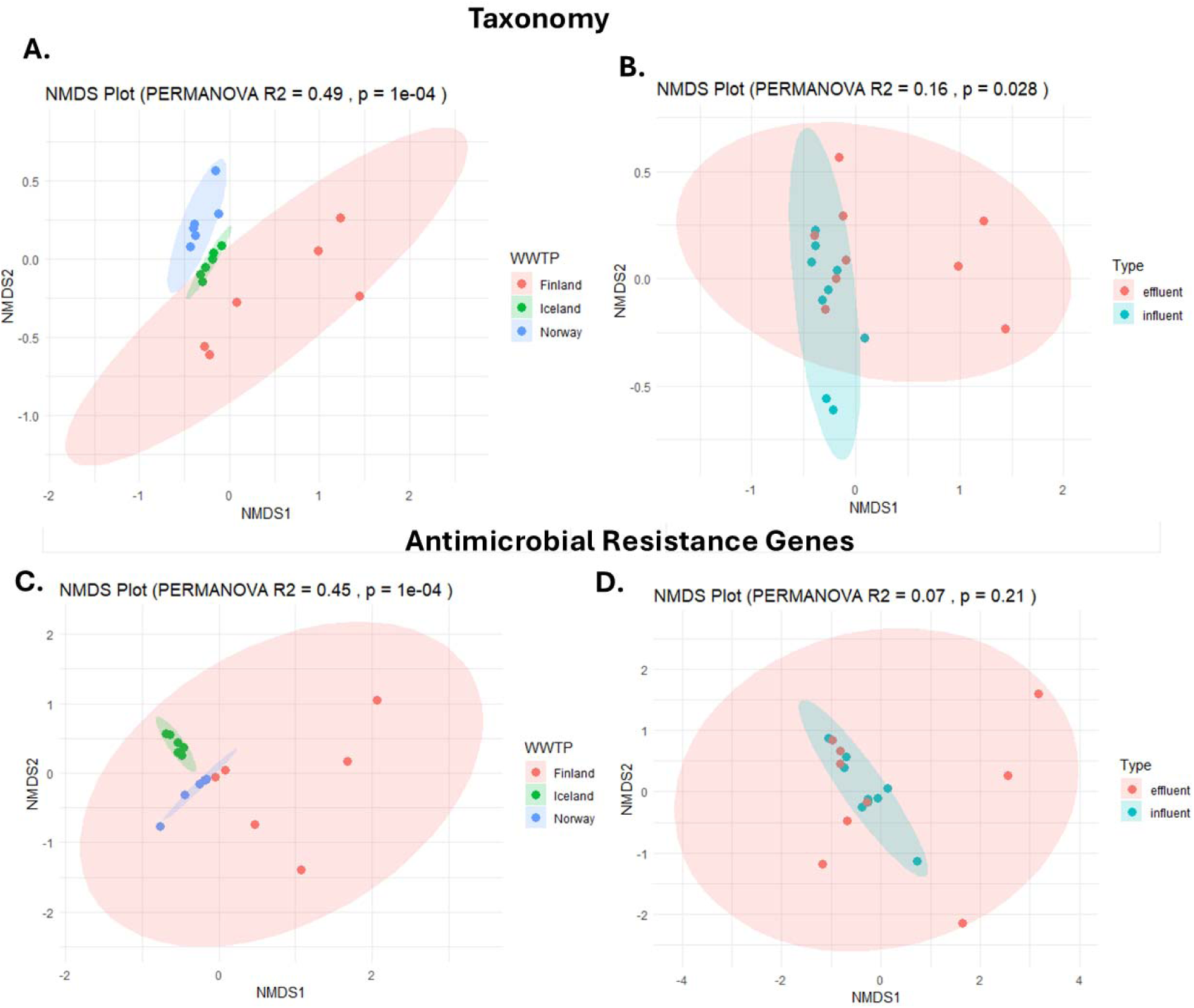
Nonmetric multidimensional scaling (NMDS) of Oxford Nanopore data across three treatment plants, influent, and effluent, using the Bray-Curtis index. (A) Bacterial taxonomic reads combining influent and effluent. (B) Influent and effluent reads combined from three treatment plants. (C) Antimicrobial resistance gene reads combining influent and effluent. (D) Antimicrobial resistance genes in influent and effluent, combined from three treatment plants. Taxonomic level for (A) and (B): genus.

Among the microbial taxonomic classes, *Gammaproteobacteria* dominated all samples, peaking in Mekjarvik influent (75.2% of reads) and lowest in Mariehamn effluent (37.5%). *Betaproteobacteria* ranked second, followed by *Alphaproteobacteria* (Figure 2). The *Alphaproteobacteria* increased post-treatment (4.3% to 9.1% in Mekjarvik, 2.3% to 3.3% in Reykjavik, and 2.5% to 7.5% in Mariehamn). All top three most dominat microbail class was *Pseudomonadota* bacterial phylum. *Bacteroidia* were second most dominant phylum, more abundant in Mekjarvik (8.3%–11.5%) and Mariehamn (8.1%, 4.1%) than in Reykjavik (3.5%–3.8%).

**Figure 2.**
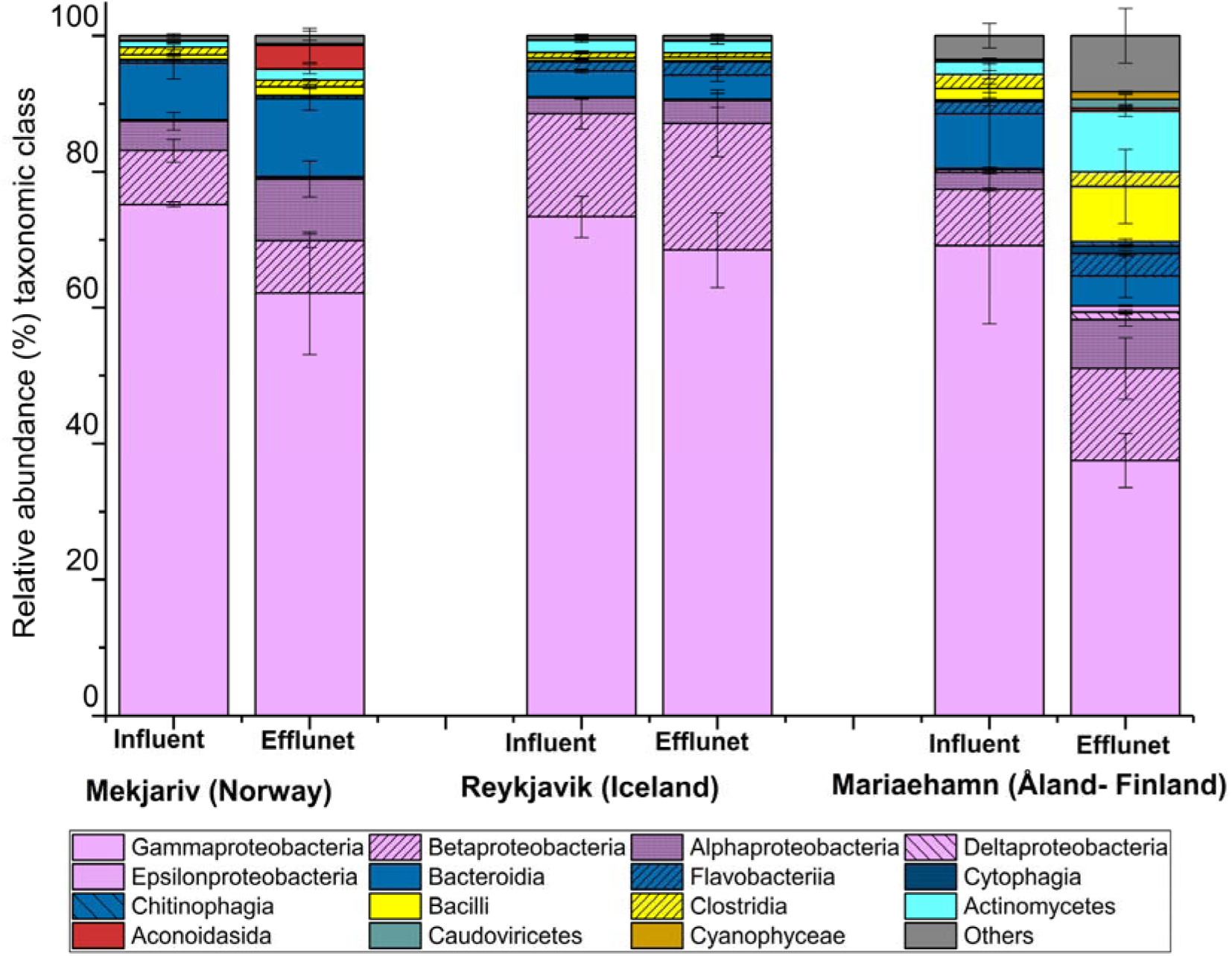
Taxonomic Composition of Microbial Communities in Wastewater Samples. Error bars represent standard deviations across replicates.

At the genus level, the top three genera in Mekjarvik influent were *Acinetobacter* (33.8%), *Aeromonas* (27.5%), and *Bacteroides* (6.9%), and also in the effluent, *Acinetobacter* (22.6%), *Aeromonas* (23.3%), and *Bacteroides* (9.6%) remained dominant. In Reykjavik influent, *Acinetobacter* (34.5%), *Aeromonas* (13.5%), and *Pseudomonas* (9.9%) were most abundant, with a similar pattern in the effluent: *Acinetobacter* (31.9%), *Aeromonas* (12.7%), and *Pseudomonas* (8.0%). Mariehamn influent was dominated by *Acinetobacter* (51.8%), *Psychrobacter* (5.7%), and *Bacteroides* (5.6%), while the effluent had *Escherichia* (15.5%), *Salmonella* (7.6%), and *Polynucleobacter* (4.8%). Among *Acinetobacter* species, the most abundant were *A. baumannii*, *A. johnsonii*, *A. lwoffii*, *A. wuhouensis*, *A. haemolyticus*, and *A. pittii*, dominating among 96 detected species. *Aeromonas;* a major genus in both Mekjarvik and Reykjavik, had total of 44 species, the most dominant were *A. caviae*, *A. veronii*, *A. hydrophila*, and *A. salmonicida*. In Reykjavik wastewater, *Pseudomonas* ranked among the top three genera, with *P. aeruginosa*, *P. fragi*, *P. lundensis*, *P. fluorescens*, *P. putida*, and *P. veronii* as the most prevalent species.

### 3.2 Detection of ARGs by Oxford Nanopore Sequencing

In total, 39,285 high-quality metagenomic ARGs sequencing reads were detected in this study. The diversity and prevlanece of ARGs distribution were differed across the three WWTPs (NMDS, PERMANOVA R² = 0.45, p < 0.001), but such diversity and distribution between influent and effluent were not significant (NMDS, PERMANOVA R² = 0.07, p = 0.21) (Figure 1). Overall, wastewater samples from Reykjavik showed the highest ARG recovery, both in influent (mean ± SD: 4,486.7 ± 1,757.5 reads) and effluent (3,757.0 ± 3,075.3 reads). These were followed by samples from Mekjarvik, with influent (2,541.7 ± 1,066.9 reads) and effluent (1,998.7 ± 1,062.8 reads). The lowest ARG recovery was observed in Mariehamn, with influent (305.7 ± 111.1 reads) and effluent (5.3 ± 4.7 reads) samples.

A total of 193 ARGs were found in our study, with the highest number (80) detected in the influent samples from a Mekjarvik sample, and the lowest in Mariehamn effluent samples (two). In general, a greater ARGs diversity was found in influent samples compared to effluent. In the influent samples, the average ARGs types were highest in Reykjavik (66.3 ± 4.1), followed by Mekjarvik (61.3 ± 14.1) and Mariehamn (18.0 ± 2.2). But, for effluent samples, the average ARG count per sample was highest in Mekjarvik (55.0 ± 16.9), followed by Reykjavik (53.3 ± 12.5), and Mariehamn (2.3 ± 0.5).

We found, *tet(39)* was the most universal ARG detected in 17/18, except in one Mariehamn effluent, followed by *tet(Q)* detected in 16, missing in two Mariehamn effluent samples. Then, *mph(E)*, *msr(E)*, and *tet(W)* were found in 15 samples except in all Mariehamn effluent samples.

In Reykjavik’s wastewater samples, a total of 141 ARGs were detected, with half appearing in only one sample and most of them were singletons. However, 21 ARGs were universally present across all influent and effluent samples. These included 11 aminoglycoside resistance genes (*aac(6’)-Im*, *aadA1*, *aadA11*, *aadA2*, *aadA6*, *ant(2’’)-Ia*, *ant(3’’)-Ia*, *aph(2’’)-Ib*, *aph(3’’)-Ib*, *aph(3’)-IIIa*, *aph(3’)-VIa*, *aph(3’)-Ia*, *aph(6)-Id*), β-lactamase genes (*bla*_ACI-1_, *bla*_AER-1_), and carbapenemase genes (*bla*_CARB-10_, *bla*_CARB-14_, *bla*_CARB-4_, and *bla*_LCR-1_). Among these, *aph(3’)-VI* (1683 ± 1258), *aph(3’)-IIIa* (870 ± 532), and *aph(3’)-Ia* (764 ± 470) exhibited the highest read prevalence.

A total of 125 ARGs were detected in Mekjarvik samples, with 50 appearing in only one sample and including 45 singletons. However, 20 ARGs were universally present across all samples, including tetracycline resistance genes (*tet(39)*, *tet(C)*, *tet(E)*, *tet(Q)*, *tet(W)*), aminoglycoside resistance genes (*ant(2’’)-Ia*, *aph(3’’)-Ib*, *aph(3’)-Via*, *aph(6)-Id*), nitroimidazole resistance genes (*nimA*, *nimD*, *nimE*), macrolides (*erm(F)*, *mph(E)*), macrolide-streptogramin B (*msr(E)*), carbapenemase (*bla*_AER-1_), β-lactamase (*bla*_OXA-129_), chloramphenicol (*cat*), quinolone (*qnrS2*), and sulfonamide (*sul2*). Notably, *mph(E)* (576 ± 308), *msr(E)* (491 ± 258), and *tet(39)* (417 ± 232) exhibited the highest ARG read counts.

A total of 36 ARGs were detected in the Mariehamn samples, half of them detected in only one sample, with 6 being singletons. No genes were universally present across all samples; however, *tet(39)* was detected in five samples, and *tet(Q)* in four samples.

#### ARG conferring different antibiotic classes

We calculated the relative abundance of ARGs conferring resistance to various antibiotic classes in influent and effluent wastewater samples from three WWTPs (Figure 3). Tetracyclines, macrolides, beta-lactams, multidrug resistance, and aminoglycosides were the most prevalent resistance classes. In Mekjarvik WWTP (Norway), with both primary and secondary treatment, showed a reduction in tetracycline, macrolide, and multidrug resistance in the effluent, suggesting partial ARG removal. However, beta-lactamase and aminoglycoside resistance slightly increased. In Reykjavik WWTP (Iceland), with only primary treatment, had a high proportion of quinolone resistance in the influent, which decreased in the effluent, while macrolide resistance remained dominant. Quinolone resistance was significantly higher in Reykjavik than in the two other WWTPs. Mariehamn WWTP (Åland-Finland), also with primary and secondary treatment, showed a notable shift in the effluent, with a higher proportion of beta-lactam and Tetracycline resistance. Overall, the beta-lactam resistance increased significantly in plants using biological activated sludge as secondary treatment, while macrolide and multidrug resistance decreased across all sites.

**Figure 3.**
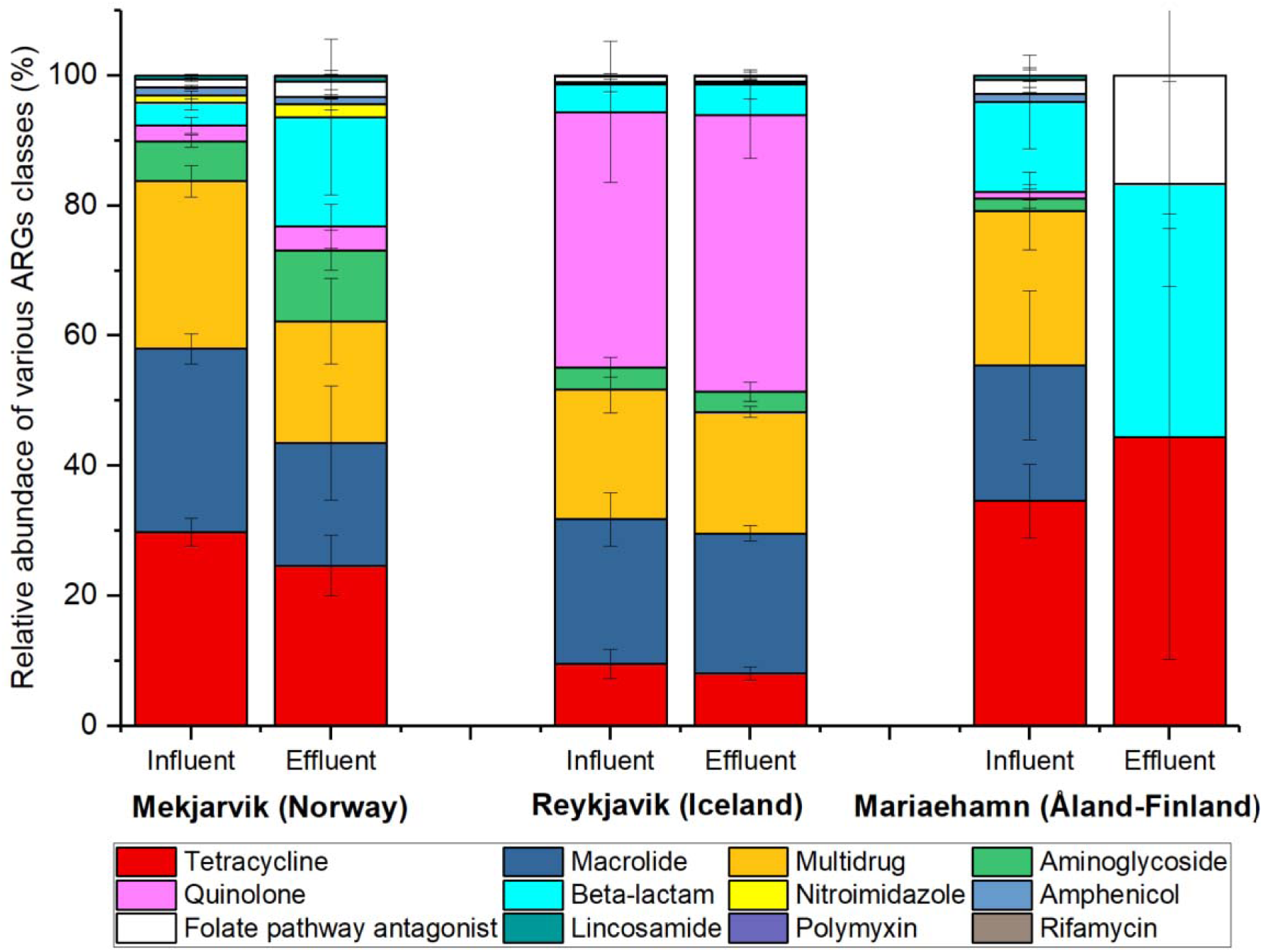
Relative abundance of ARGs conferring resistance to various antibiotic classes detected in influent and effluent of three wastewater treatment plants using Oxford-Nanopore high-throughput metagenomic sequencing of total nucleic acids.

**Figure 4.**
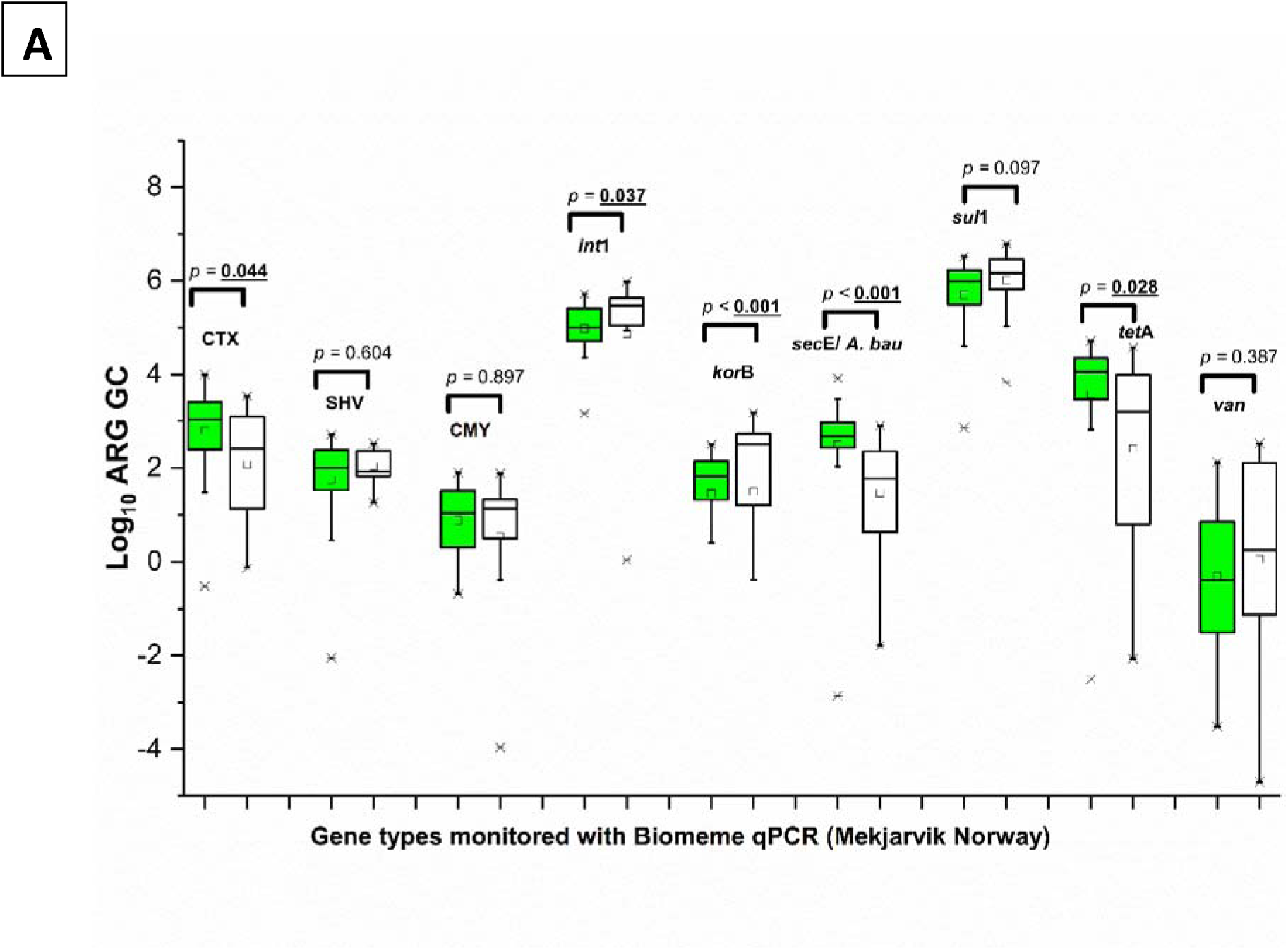
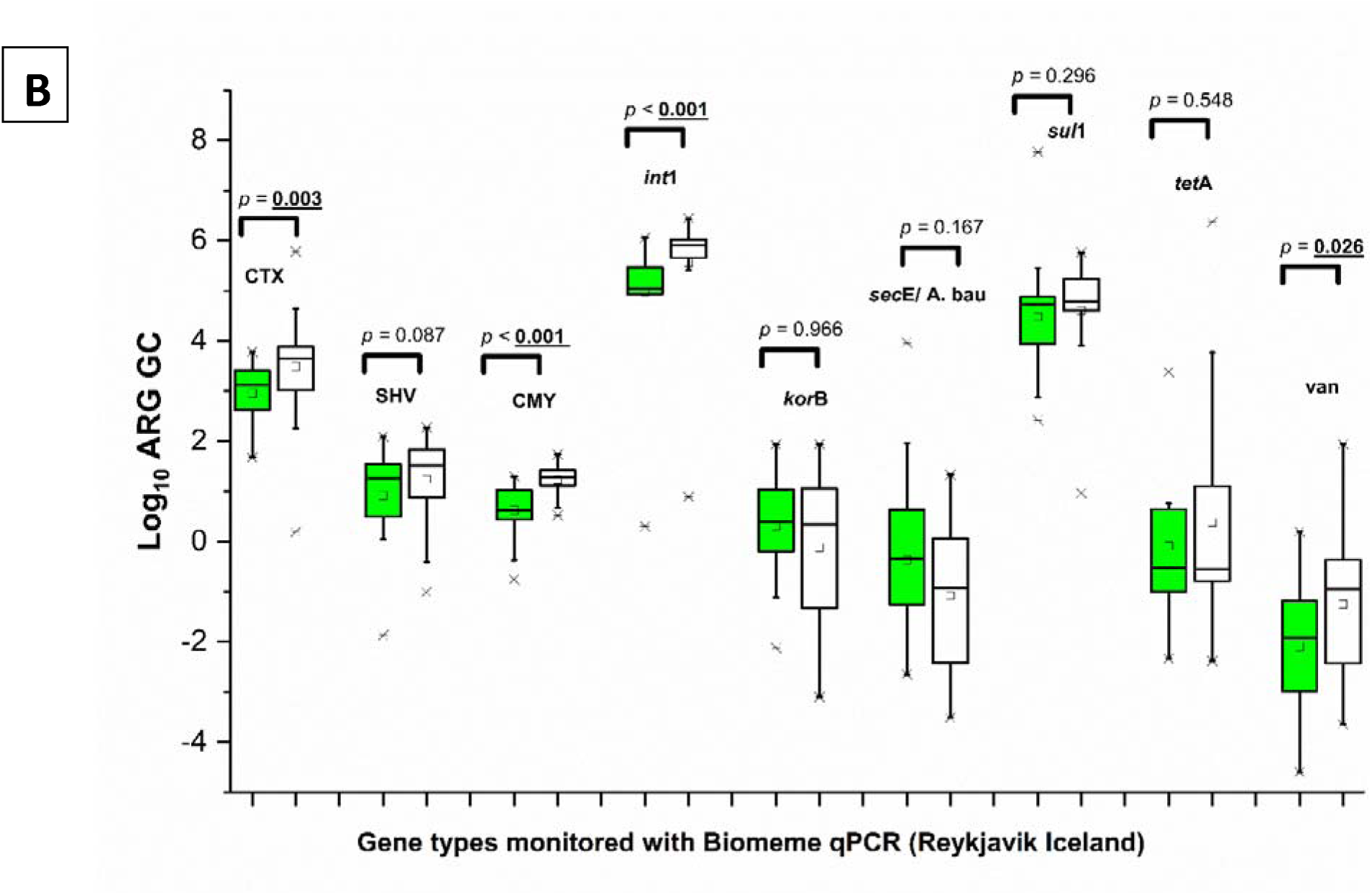
Box-plot representation of gene copy numbers for various targeted ARGs using the Biomeme qPCR method. The black line indicates the median, the square marker represents the arithmetic mean, and the box’s upper and lower edges correspond to the third and first quartiles, respectively. (A) Mekjarvik (Norway) (B) Reykjavik (Iceland). The Mann-Whitney U test was used to test for significance.

**Figure 5.**
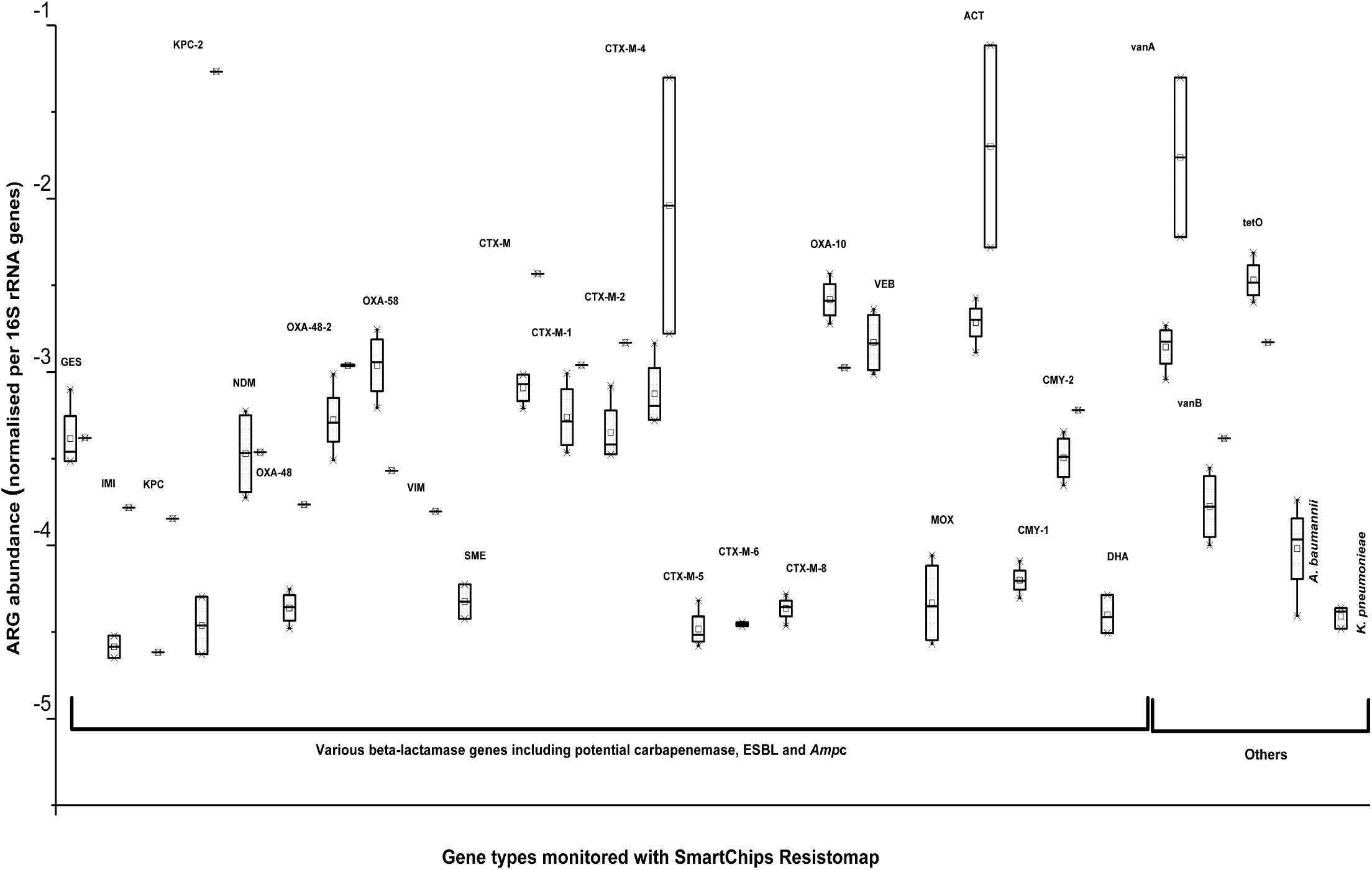
Abundance of various antimicrobial and taxonomic genes in influent and effluent samples collected from Mariehamn. (Gray represents influent and white represents effluent).

Table S4 shows the major attenuated and amplified genes in each sample in three WWTP. The number of attenuated ARGs consistently exceeded amplified ARGs, except in two samples. In July at Mekjarvik, the pattern reversed, with many aminoglycoside, macrolide, and tetracycline genes amplified, and only two ESBL and carbapenem genes attenuated.

In May at the Reykjavik WWTP in Iceland, aminoglycoside, β-lactam, macrolide, and tetracycline genes were largely amplified, while others in the same classes were attenuated. In May, ten ARGs—primarily conferring resistance to β-lactams, carbapenems, aminoglycosides, macrolides, chloramphenicol, and sulfonamides—showed higher read counts in Reykjavik effluent than in influent samples.

#### Plasmid types and ARGs

A total of 13 plasmid types associated with ARGs were detected with ONP, majors were *Col440, IncQ1, IncQ2,* and *ColRNAI, and IncP6,* carrying genes that confer multidrug resistance, were the most prevalent, found in many wastewater samples nearly all WWTPs highlighting their role in HGT (Table 2). Plasmid-associated ARGs conferred resistance to multiple antibiotic classes, with high prevalence in quinolones, tetracyclines, beta-lactams, aminoglycosides, macrolides, and sulfonamides across all WWTPs, regardless of the presence or absence of secondary treatment. Plasmid-associated ARG diversity was higher in Mekjarvik (with secondary treatment), spanning all antibiotic classes (e.g., *aph*(3’’)-Ib, *bla*_TEM_, *qnr*, *aph*, *sul*2, *tet*(C)). Reykjavik (only with primary treatment) predominantly had quinolone resistance genes (*qnr*-variants) but also *aph*(3’’)-Ib, bla_OXA_, and *tet*(C) (Table 2). Despite having secondary operation unit, the Mariehamn WWTP had fewer ARGs, conferring resistance to macrolides, lincosamides, and streptogramins (Table 2). Among plasmid-mediated ARGs, *qnr*B19, *qnr*S2 and *tet*(C) were among the most widely distributed genes in majority of samples in all WWTPs, in both influent and effluent.

**Table 2:**
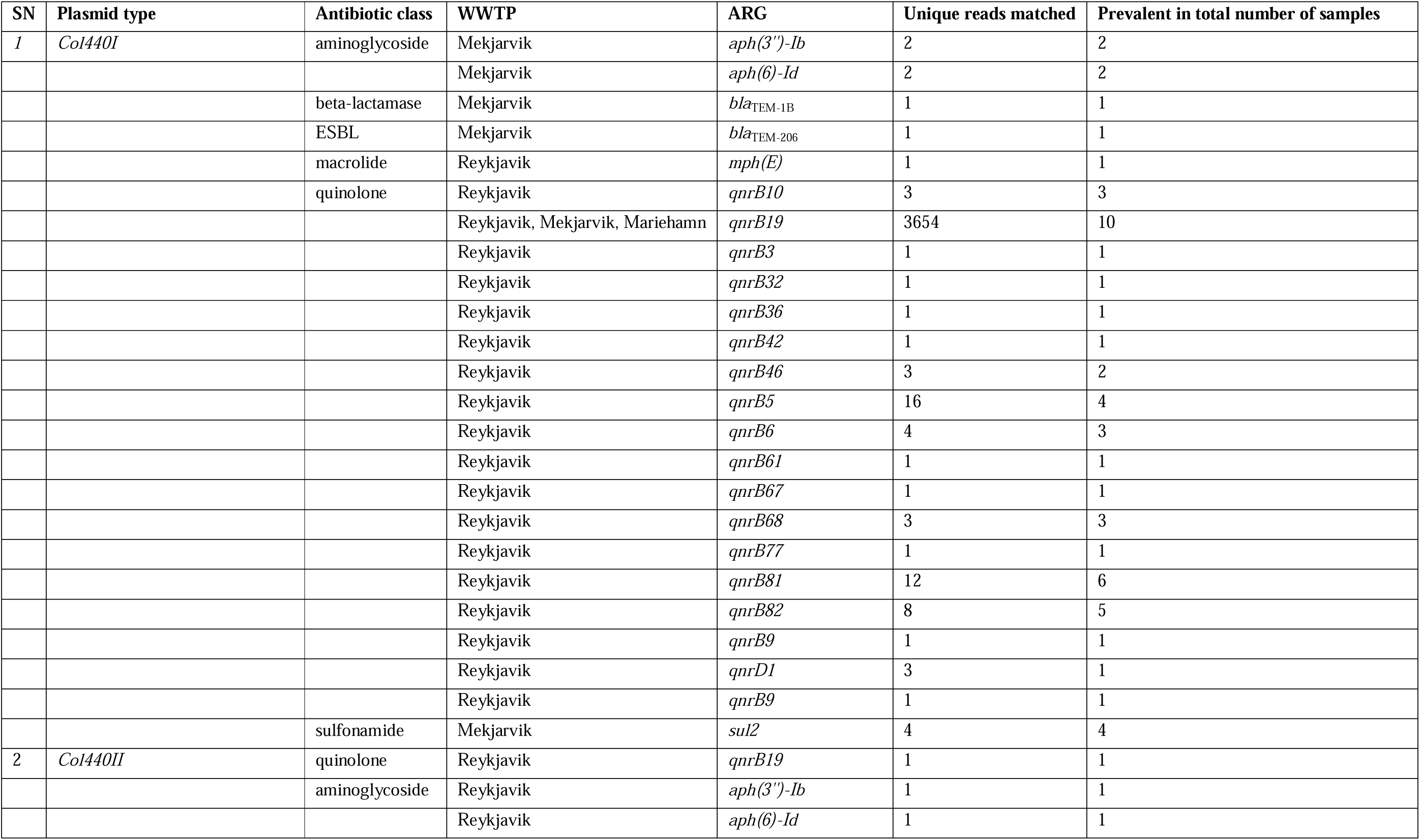
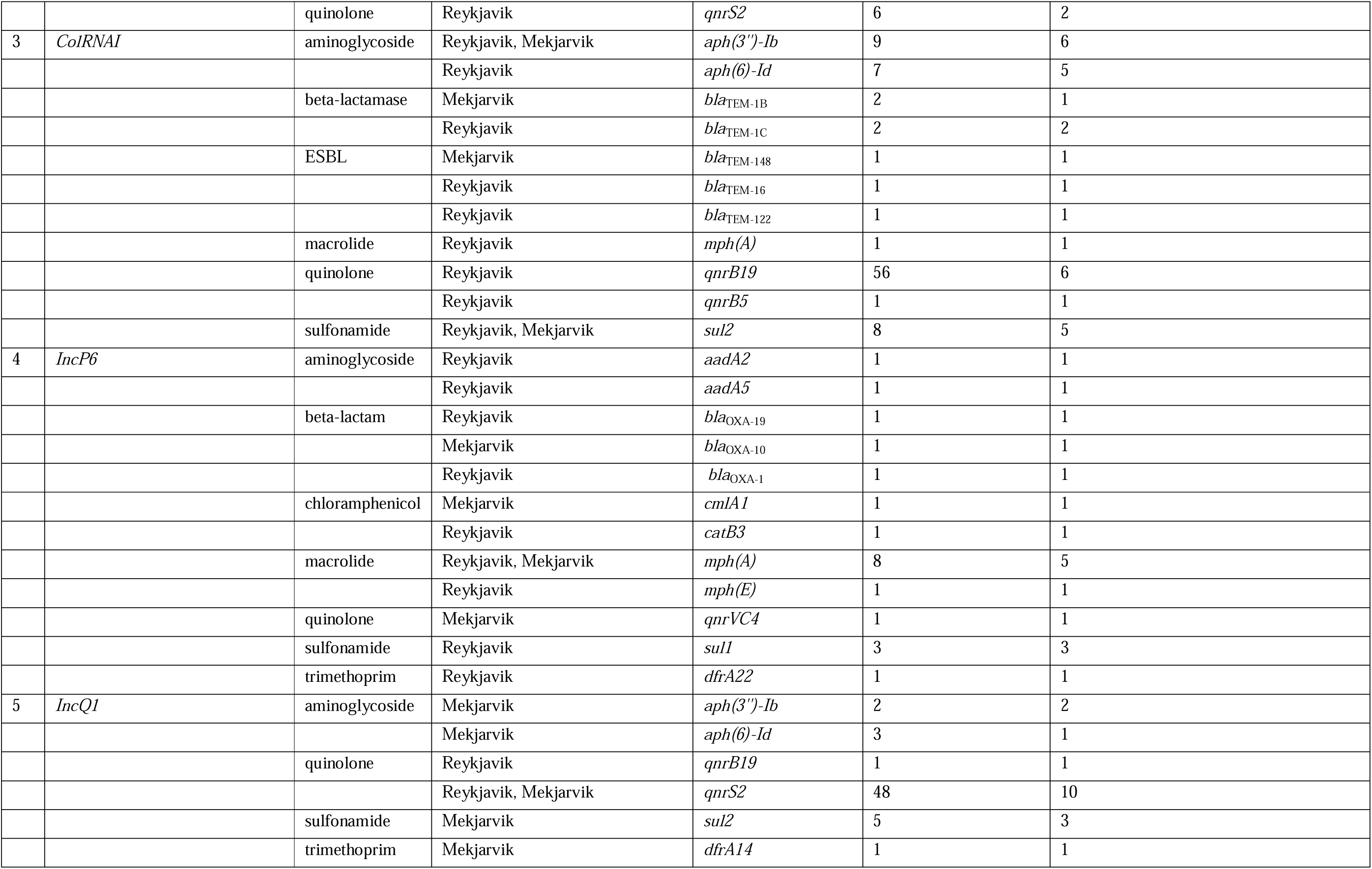
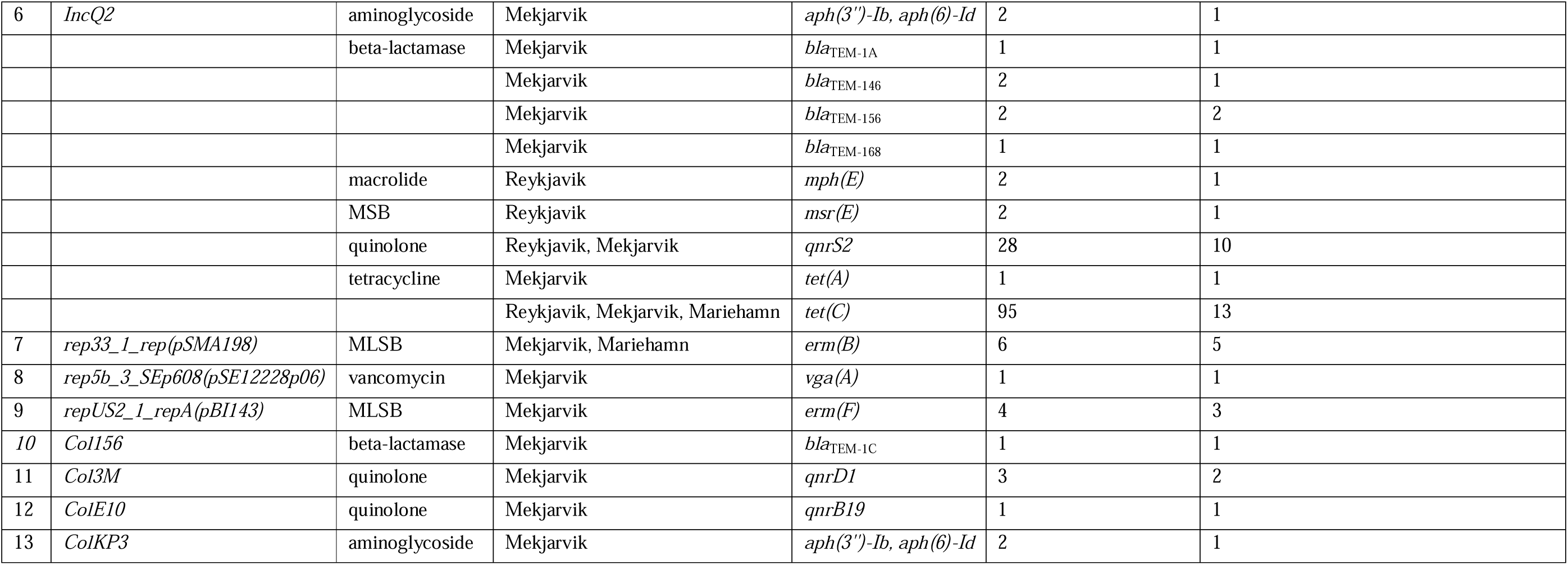
Prevalence of Plasmids and Antimicrobial Resistance Genes in Wastewater Treatment Plants.

### 3.3 Detection of ARGs with qPCR Methods

Prevalence of ARGs in the Mekjarvik (Norway) and Reykjavik (Iceland) samples was monitored also using a Biomeme qPCR platforms. ARGs were monitored across 11 episodes in Reykjavik and nine episodes in Mekjavik WWTPs. Among the 9 targeted ARGs, the prevalence of the *int1* and *sul1* genes exhibited the highest prevalence, while the *van* gene had the lowest prevalence in both WWTPs. Interestingly, in the Reykjavik WWTP, the gene copies of *bla*_CTX-M,_ *bla*_CMY_, *int1*, and *van* were significantly higher in the effluent compared to the influent. While, in the Mekjarvik WWTP, *int1* and *korB* showed significantly higher gene copies in the effluent than in the influent. Meanwhile, *bla*_CTX-M_, *secE*, and *tetA* exhibited significantly higher gene copies in the influent than in the effluent.

Prevalence of ARGs in the Mariehamn samples was monitored using a SmartChip qPCR Resistomap system, four episodes in influent and three episodes in effluent samples. Among the 13 targeted potential carbapenemase genes-*bla*_GES_, *bla*_NDM_, *bla*_OXA48_, and *bla*_OXA58_ were detected in all influent samples, but only in one of the three effluent samples. Genes: *bla*_IMI_, *bla*_KPC_2_, and *bla*_SME_ were detected in two out of the four influent samples, and these genes were also detected in one effluent samples, but *bla*_SME_ was not detected in the effluent samples. *bla*_VIM_ was found in only one effluent sample and was not detected in any influent samples. The *bla*_KPC_ was detected in one influent and one effluent sample. ARGs: *bla*_IMP_1_, *bla*_KPC_3_, and *bla*_OXA23_ were not detected in any influent or effluent samples in this study.

Among targeted nine ESBL genes, seven were *bla*_CTX-M_ types along with *bla*_OXA10_1_ and *bla*_VEB_. All ESBL genes were detected in all influent samples, except for *bla*_CTX-M_6_, which was found in only in two influent samples. In effluent samples, *bla*_CTX-M_, *bla*_CTX-M_1_, *bla*_CTX-M_2_, and *bla*_OXA10_ were detected in one sample, while *bla*_CTX-M_4_ was found in two samples. *bla*_CTX-M_5_, *bla*_CTX-M_6_, and *bla*_CTX-M_8_ were not detected in effluent samples. Four of the five targeted *amp*C β-lactamase genes (*bla*_DHA_, *bla*_CMY_1_, *bla*_CMY2_, *bla*_MOX_*/bla*_CMY_) were detected in all four influent samples, with *bla*_DHA_ found in three. In effluent samples, *bla*_ACT_ was detected in two samples, and *bla*_CMY2_ in one, while the other three were undetected.

Vancomycin resistance genes (*van*A and *van*B) and the tetracycline resistance *tet*O were abundant in all influent samples. The *van*A was detected in two effluent samples, while *van*B and *tet*O were each found in only one effluent sample. The methicillin resistance *mec*A was not detected in any influent or effluent samples. The *crAss*56 gene targeting crAssphage was present in all wastewater samples. Gene specific to *A. baumannii* was found in all influent samples, while those for *K. pneumoniae* appeared in three influent samples, but both these genes were absent in all effluent samples. No *Staphylococci* spp. genes were detected in any samples.

Most ARGs monitored in this study showed clear higher detection rates in influent than in effluent. However, carbapenemase genes (*bla*_IMI_, *bla*_KPC_, *bla*_OXA-48_), ESBL genes (*bla*_CTX-M_, *bla*_CTX-M_1_, *bla*_CTX-M_2_, and *bla*_CTX-M_9_), *amp*C β-lactamase genes (*bla*_CMY2_, and *bla*_ACT_) and vancomycin resistance genes (*van*A and *van*B) showed higher relative gene copies in effluent than in influent.

#### Prevalence of metal stress genes and possible co-selection pressure

With ONP metagenomic sequencing, a total of 966 metal stress reads were recorded, with the majority related to mercury (64.9%), followed by multimetal (23.7%), copper (6.4%), biocide and metal (2%), nickel (1.8%), and others, including zinc, tellurium, chromium, cadmium, and arsenic. As shown in Figure S2, influent samples had clear higher metal stress reads than effluent samples. Among the three WWTPs, most reads were recorded in the Mekjarvik and Reykjavik treatment plants. Of the total reads, 722 (74.7%) were associated with heavy metal resistance proteins, 222 (23%) with heavy metal stress regulator proteins, and 17 (1.8%) with ABC efflux pumps. The remaining ∼0.5% were linked to RND efflux pumps, ABC efflux regulators, MFS efflux pumps, and MFS efflux regulators. Notably, most reads conferring nickel resistance were associated with ABC efflux pumps. Mercury, multimetal, and copper-related reads were universally detected across all WWTPs. However, biocide and metal-related reads were predominantly found in Reykjavik and Mariehamn samples, while chromium-related reads were mostly observed in Reykjavik samples.

**Figure S2.**
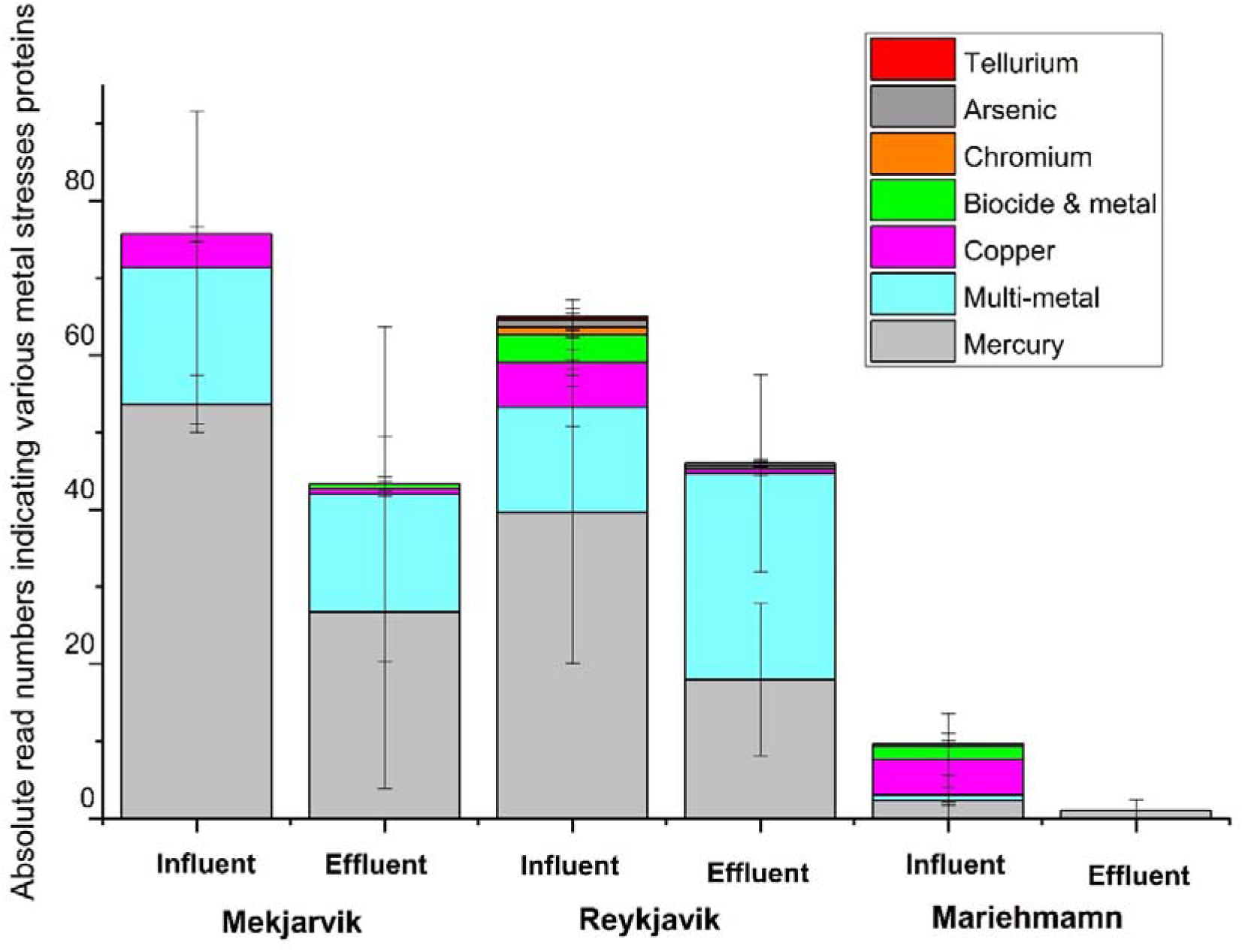
Distribution of metal resistance genes in influent and effluent samples from various WWTPs. Error bars represent standard deviation from three sampling events.

### 3.4 Prevalence of antibiotic residues

Of the 45 antibiotics screened in this study (Table S1), only five were detected with sufficient confidence to be reported (Figure 6). We detected majorly only sulfamethoxazole, sulfapyridine, azithromycin, ciprofloxacin, and ofloxacin, with clear high levels of sulfamethoxazole and sulfapyridine in both WWTPs. The concentration of azithromycin, ciprofloxacin, and ofloxacin remained mostly below detection limits at both sites. The concentration of detected antibiotics was varied over samples, in Mekjarvik, the level peaked at ∼2000 ng/L in April, with most samples around ∼600 ng/L; lower levels (∼200 ng/L) were seen in July, September, and October. Reykjavík samples consistently showed lower antibiotic levels, typically around 200 ng/L, with a peak of 300 ng/L in December. Mekjarvik showed significant monthly variation, while Reykjavík remained relativly stable across monitored episodes (Nov-Jan).

**Figure 6.**
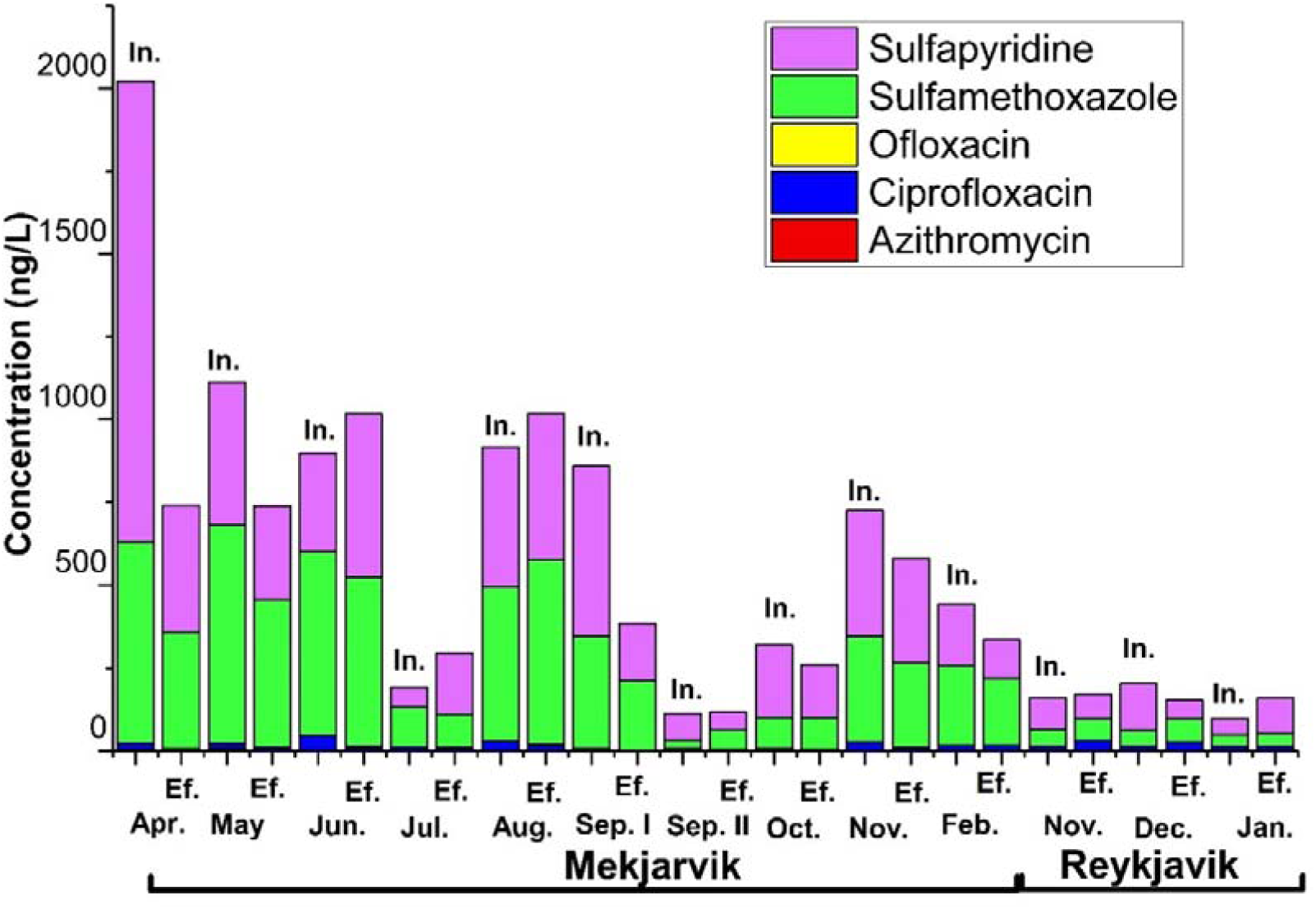
Prevalence of antibiotic residues in Mekjarvik and Reykjavik wastewater treatment plant during the study period. (In. = influent, Ef. = effluent)

### 3.5 Prevalence of microplastics

Microplastic (MPs) particles were monitored only at Mekjarvik and Reykjavik WWTPs. The highest count was in Reykjavik influent (8200 MPs/m³), followed by Mekjarvik influent (5900 MPs/m³). Reykjavik effluent had 1100 MPs/m³, while Mekjarvik effluent had the lowest at 750 MPs/m³ (Figure 7). Polyethylene accounted for about one-third of total MPs in Reykjavik influent and effluent and in Mekjarvik effluent. However, in Mekjarvik influent, nearly half of the MPs were Polypropylene. Polystyrene made up roughly one-fifth of MPs in Reykjavik influent wastewater. Other major MP distributions are shown in Figure 7.

**Figure 7.**
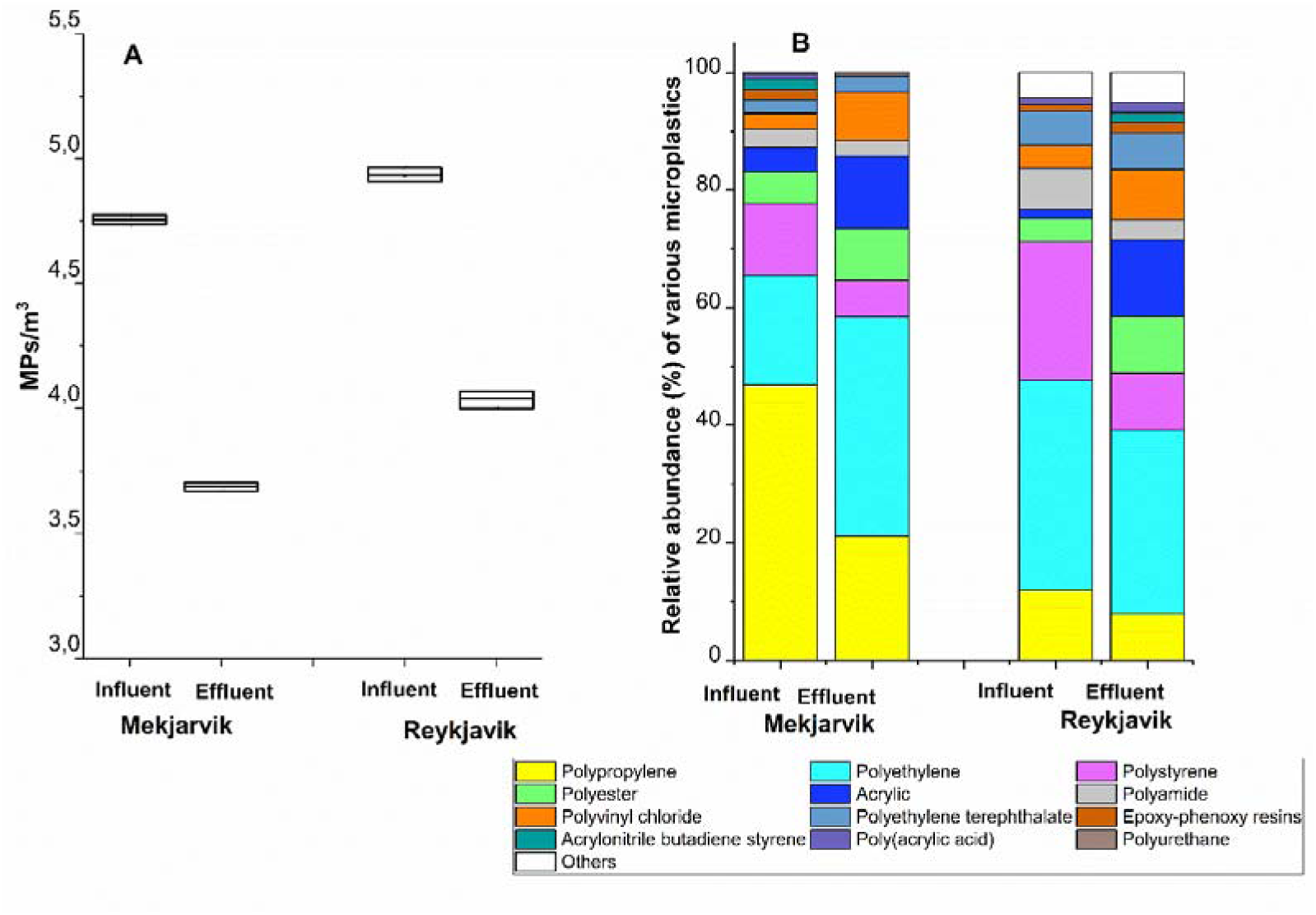
Variation in MP count and types in influent and effluent from Mekjarvik and Reykjavik WWTPs. (A) MP count. (B) MP types.

The distribution of MPs size varied across wastewater types and locations. Mekjarvik influent samples predominantly contained the 101-150 µm fraction, followed by the 151-200 µm and 201-300 µm sizes. In Reykjavik influent, the 50-100 µm and 101-150 µm sizes were also prominent. In both effluent samples, fine MPs increased while larger particles decreased after treatment (Figure S3).

**Figure S3.**
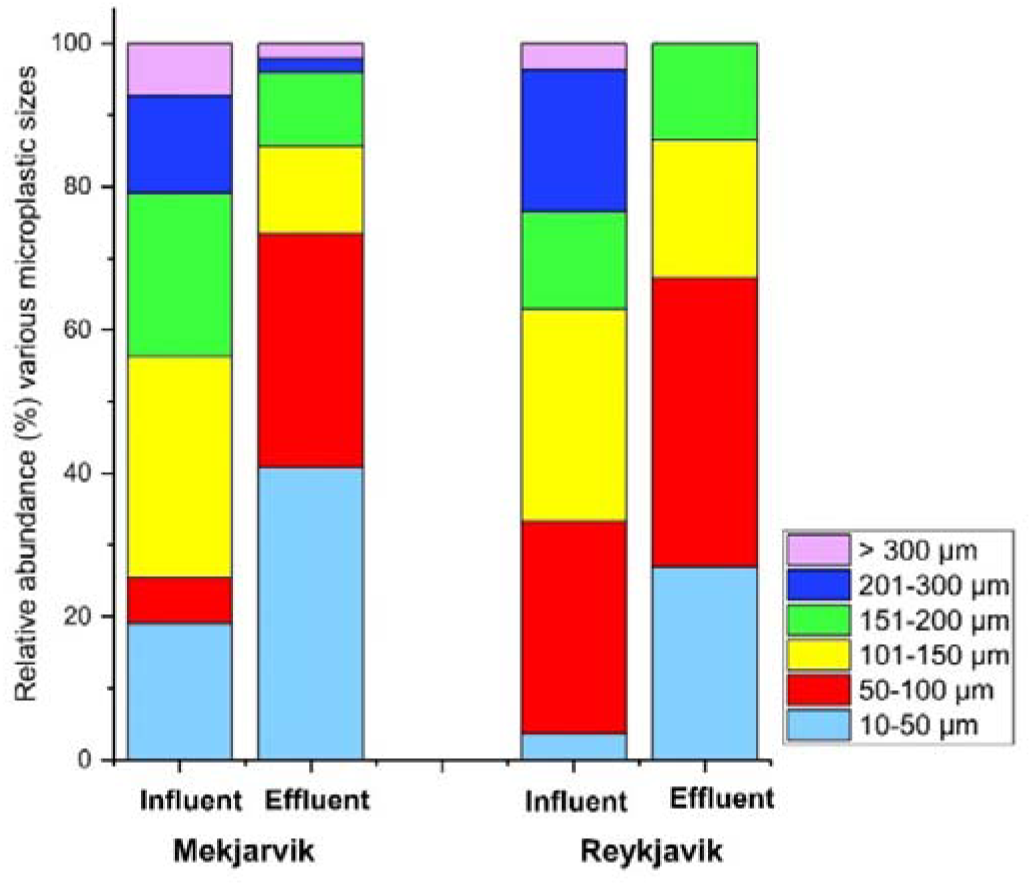
Relative abundance of microplastic sizes in influent and effluent at Mekjarvik and Reykjavik WWTPs.

### 3.6 SOS Response: Bacterial DNA Damage Repair

In Mekjarvik (with secondary treatment unit), SOS *E. coli* fluorescence levels were significantly higher in influent than effluent samples in May and June. However, in July, effluent levels exceeded influent levels. Average fluorescence levels (± SE) for influent samples in May, June, and July were 4.09 ± 0.06, 4.25 ± 0.06, and 3.98 ± 0.02, respectively, while effluent averages were 3.57 ± 0.07, 3.74 ± 0.07, and 4.16 ± 0.02. In Reykjavik (only with primary treatment unit and wastewater dilution using home-heated water), showed a higher counts in influent only in November, with effluent levels exceeding influent levels in December and January, among monitored in three consecutive months. Average influent fluorescence levels (± SE) were 4.01 ± 0.04, 3.84 ± 0.06, and 3.76 ± 0.05, while effluent averages were 3.75 ± 0.09, 3.09 ± 0.04, and 3.75 ± 0.10, respectively across November, December, and January (Figure 8).

**Figure 8.**
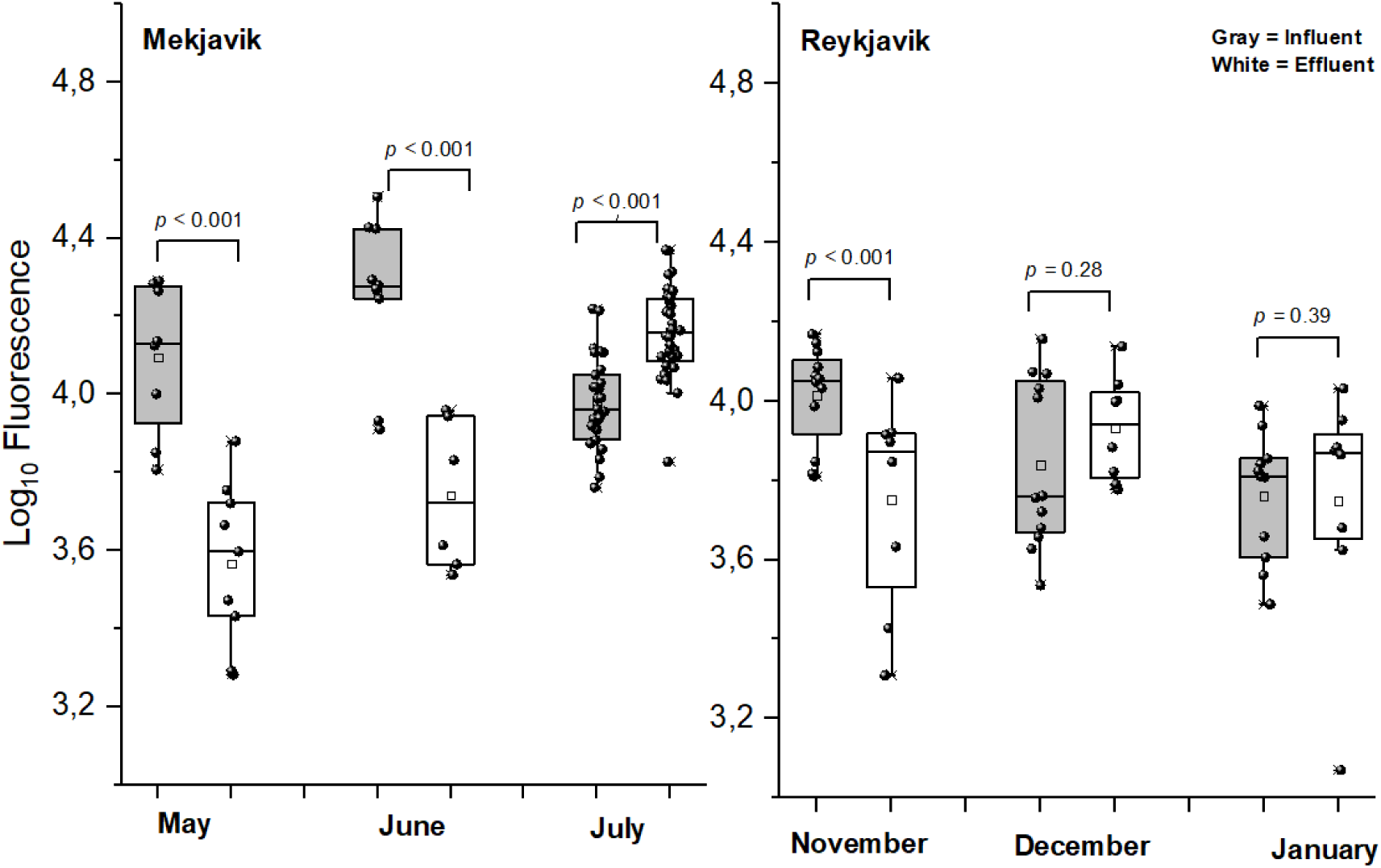
SOS *E. coli* fluorescence levels in influent and effluent samples from Mekjarvik and Reykjavik WWTPs across different sampling months. Each dot represents measurement of sample replicates from each monitoring event. The Mann-Whitney U test was used to test for significance.

**Figure S4.**
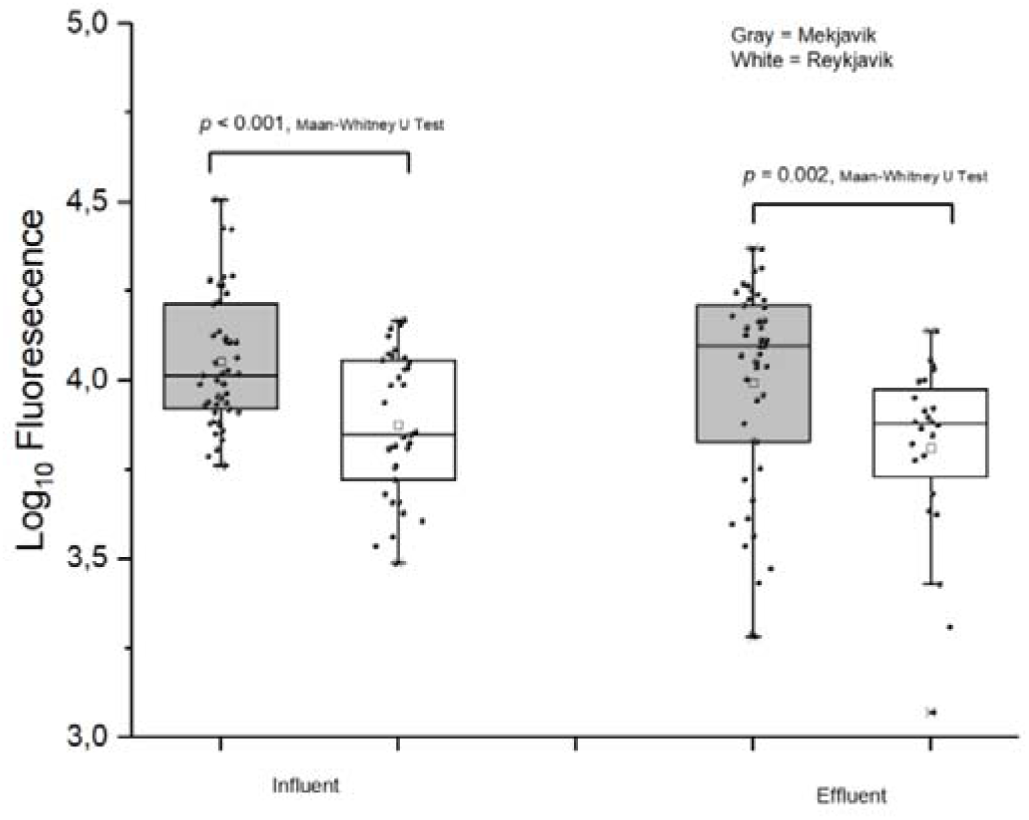
Comparison of SOS *E. coli* fluorescence levels in influent and effluent samples from Mekjarvik and Reykjavik WWTPs, based on pooled samples from all three months. Each dot represents a measurement from each samplings including sample replicates. The Mann-Whitney U test was used to test for significance.

## 4. Discussion

This study monitored ARGs in influent and effluent at three Nordic WWTPs (Reykjavik, Iceland; Mekjarvik, Norway; and Mariehamn, Åland-Finland), but antibiotic residues and MPs monitoring were only feasible in Reykjavik and Mekjarvik. The presence of plasmid-mediated ARGs, metal stress genes, and opportunistic pathogens (e.g. *A. baumannii, P. aeruginosa, Aeromonas spp., Bacteroides spp., E. coli*, *Salmonella spp.*) in both influent and effluent highlights WWTPs’ potential role in ARB pathogen dissemination. *A. baumannii*, and *P. aeruginosa* are nosocomial pathogens with environmental sources, are leading cause of ARB-related deaths globally ^45^, and are on the WHO critical ARB list, requiring urgent development of alternative treatments ^5^.

The presence of plasmid-mediated ARGs, metal stress genes, and opportunistic pathogens in wastewater influent underscores the high potential for HGT, co-selection, and MDR development during their flow from households to WWTPs facilitates, complicating interpretation of WS results, and effort to linking AMR in wastewater and their prevalence at population-level ^3–5^. Micropollutants, including pharmaceuticals, MPs and heavy metals, accelerate co-selection, as metal resistance genes often co-occur with ARGs, enabling resistance even without antibiotics ^46^. The spread of ARGs in wastewater is influenced by ecological factors, such as environmental conditions (temperature, pH, nutrients, oxygen), which influence microbial interactions and ARG persistence. In WWTPs, microbial interaction and diversity impacts HGT mechanisms (conjugation, transduction, transformation), facilitating ARG transfer ^47^. Biofilms and high bacterial densities in secondary treatment enhance gene exchange, promoting AMR dissemination. While ARG levels generally decrease (potentially due to reduced bacterial loads), many persist and may increase relative to total bacterial reduction. Our SOS-*E. coli* analysis, assessing genotoxicity, DNA damage, and stress responses of *E. coli* strains exposed to environmental contaminants (including antibiotics and pollutants), reduced SOS activity during the treatment process. In May and June, WWTPs in Mekjavik successfully removed a significant proportion of bacterial stressors (e.g., antibiotics), whereas the Reykjavik WWTP was less effective. Well-operated WWTPs with both primary and secondary treatment units are more effective at removing bacterial stressors than those with only primary treatment. However, overall, wastewater from Mekjavik contained a higher quantity of stressors than that from Reykjavik, regardless of whether in the influent or effluent.

Interestingly, some ARGs were amplified in Reykjavik WWTP with only primary treatment. This may be due to elevated sewage temperatures, as wastewater in Reykjavik is collected in house basements, mechanically screened, and mixed with heated household water before releasing them into ocean. The higher temperature in the system could enhance HGT of ARGs, as the biological activities of enteric pathogens are optimum in mesophilic range enhancing HGT of ARGs. Previous studies show that in WWTPs, ARG abundance and *intI*1 gene prevalence correlate positively with effluent temperature, influencing microbial communities and ARG proliferation ^48,49^. The optimal temperature range for biological reactions is 15–35 °C, where increased temperatures promote bacterial growth and ARG spread ^49^. In contrast, lower temperatures may reduce acetylation enzyme activity, which is crucial for transcriptional regulation and microbial growth ^50^.

Detection of ESBL and carbapenemase genes (*bla*_TEM_*, bla*_OXA_*, bla*_KPC_*, bla*_GES_) signals resistance to third-generation cephalosporins and carbapenemase, while MLSB resistance genes (*ermB, ermF*) indicate macrolide resistance ^51,52^. ARGs in effluent pose pollution risks to surface waters, making effluent monitoring vital for assessing public health threats.

Although WWTPs reduce ARG prevalence, dominant ARGs often persist post-treatment, highlighting the inadequacy of current processes and the need for strategies to mitigate AMR spread ^3,4^. Site variations likely reflect regional antibiotic use, treatment technologies, and environmental factors influencing ARG persistence ^27^.

Our findings confirm the link between regional antibiotic consumption and ARG prevalence, reinforcing WS as a key tool for monitoring ARGs at the population level. The higher fluoroquinolone/ciprofloxacin, tetracycline, and β-lactam consumption in Iceland compared to Finland and Norway, was well mirrored with an increased prevalence of corresponding ARGs in Reykjavik wastewater ^27,53^. Tetracyclines, widely used in human and veterinary medicine in the Nordic region ^53^, along with macrolides for respiratory and skin infections, and quinolones for urinary and respiratory tract infections, contribute to the spread of resistance ^53^.

Resistance to β-lactams, particularly carbapenems, poses a major threat to clinical care, public health, and animal health ^5,54^. Carbapenems are critical for treating severe infections caused by ESBL-positive pathogens ^55^, but their overuse fosters the proliferation of multidrug-resistant pathogens, making infections increasingly difficult to treat ^56^. This resistance is rising in Nordic countries, often imported by travelers and hospitalizations abroad ^27^. In Reykjavik, broad-spectrum antibiotics, including azithromycin, exert selective pressure on microbes, increasing the potential for AMR due to excretion into the environment. This highlights the urgent need for careful antibiotic prescription to mitigate resistance risks in both clinical and environmental settings.

This study piloted portable devices, including the Biomeme Franklin qPCR device and the Oxford Nanopore MinION sequencing device, for qualitative and semi-quantitative ARG assessment. These tools were tested for monitoring ARGs in WWTP influent and effluent, addressing the global need for early AMR warning via WS and evaluating public health risks associated with wastewater effluent discharge into surface waters. Our toolbox with portable solutions to monitor WWTP influent and effluent combined with semi-autonomous data analysis and near-real-time data visualizations can act as an effective early warning system for AMR spread in population level and effective tool for accessing the efficiency of wastewater treatment.

We observed consistency between qPCR and metagenomics in measuring ARGs, with β-lactamase and carbapenemase genes consistently amplified during the biological treatment process. However, due to method throughput, broader comparisons were not possible.

Perhaps it would be important to consider that the whole genome amplification (WGA) used prior metagenomics might introduce some bias on the abundance of genes. Still, metagenomics offers a complete catalog of ARGs in wastewater, while qPCR provides targeted results based on selected genes.

### Antibiotic residue in wastewater: impact of community usage and environmental fate

The detection of antibiotic residues in wastewater is not only determined by their use in community, but also by their fate and decay rate in the environment. Out of 45 antibiotics screened, only five were detected with sufficient confidence. Despite tetracyclines (J01A) and beta-lactam penicillins (J01C) being the most commonly used antibiotics in Norway and Iceland ^53,57^. Other major antibiotics used in Reykjavik were macrolides/azithromycin (11.4%), tetracyclines/doxycycline (7.8%), pivmecillinam (7.4%), cephalosporins/cephalexin (4.7%), sulfamethoxazole and trimethoprim (2.7%), nitrofurantoin (2.4%), trimethoprim (2.3%), fluoroquinolones/ciprofloxacin (2%), and phenoxymethylpenicillin (1.8%) ^57^. The absence of many of these antibiotics in wastewater may suggest their rapid decay in sewage. Each antibiotic has unique mechanism absorbing in human body and fate and decay rate in environment. For instance, penicillin is unstable under environmental conditions, and tetracyclines are also sensitive to pH levels or temperatures^58^. Many of these antibiotics may have been diluted in the sewer system before sampling, falling below the detection limits of analytical methods. A literature review reported, sulfonamides are among the most commonly detected antibiotics in wastewater ^59^.

Among the most detected antibiotics, Sulfamethoxazole, often combined with trimethoprim, and sulfapyridine, frequently paired with sulfasalazine, are sulfonamides that inhibit folic acid synthesis, essential for bacterial growth, and are widely used to treat urinary and respiratory infections and certain inflammatory conditions. Azithromycin, a macrolide effective against respiratory, skin, and sexually transmitted infections, inhibits bacterial protein synthesis. Ciprofloxacin and ofloxacin, both fluoroquinolones, disrupt bacterial DNA replication and are used for urinary, respiratory, and skin infections.

### Well-operated WWTPs remove larger size MPs

This study demonstrates that a well-operated WWTP can eliminate a large fraction of MPs, but may not be as effective for smaller-sized MPs. This finding is consistent with earlier research, which reported that full-scale WWTPs with both chemical and biological treatment (in Sweden and Finland) removed more than 99.7% of MP particles ≥300 µm in the influent during the treatment process ^29^. However, this level of removal was not observed in a treatment plant with only a primary treatment unit (Iceland) ^29^. Polyethylene, widely used in packaging like plastic bags and bottles, is widely used plastics that easily break down into MPS in aquatic environments. Polyamide (nylon) is common in textiles, automotive parts, and packaging. Polystyrene is often found in disposable cutlery, food containers, and insulation, while Polypropylene is prevalent in food packaging, automotive parts, and textiles. The role of MPs on ARGs dynamics in wastewater needs more studies.

## 5. Conclusion

Our findings show a concordance between ARG prevalence in wastewater and broad antibiotic consumption patterns, confirming WS as an effective tool for monitoring ARGs at the population level. Portable devices used in this study provide valuable data on AMR and ARB presence. Combining quantitative (Biomeme) and qualitative (MinION Nanopore) analysis of environmental DNA, without heavy equipment, offers an optimal solution for WS in distance isolated populations. This study may contribute to the implementation of the EU’s Urban Wastewater Treatment Directive (UWWTD) ^15^ and the development of a unified ARG WBS system in Nordic countries ^27^.

Plasmid-associated ARGs, particularly multidrug resistance, were prevalent across all WWTPs, with Mekjarvik showing the highest plasmid-associated ARG diversity, followed by Reykjavik and Mariehamn. Ecological factors and treatment processes shape ARG diversity. Quinolone, tetracycline, macrolide, and beta-lactam resistance were widespread. Mekjarvik showed reduced tetracycline, macrolide, and multidrug resistance, while beta-lactamase and aminoglycoside resistance slightly increased. Reykjavik exhibited higher quinolone resistance, which decreased in effluent, while Mariehamn showed shifts in resistance to beta-lactams and tetracyclines in effluent. *Tet*, *Mph(E)*, and *msr(E)* were the most detected genes. Comparatively variation on microbial communities and ARG prevalence between influent and effluent were minimal.

Five antibiotics—sulfamethoxazole, sulfapyridine, azithromycin, ciprofloxacin, and ofloxacin—were detected, with higher levels of sulfamethoxazole and sulfapyridine in Mekjarvik and Reykjavik samples. Up to ∼ 90% of MPs were reduced during treatment, with most remaining as fine MPs. These results suggest that treatment processes, especially those with both primary and secondary units, alter micropollutants, pathogens and ARG profiles but do not eliminate them, facilitating horizontal gene transfer of ARGs among bacteria. The detection of diverse ARGs, plasmids, and genetic markers of critical pathogens like *A. baumannii*, *P. aeruginosa*, *E. coli*, and *Salmonella* in wastewater effluent presents a significant environmental challenge and emerging pollutant for recipient waters.

## DECLARATIONS

### CRediT authorship contribution statement

AdK conceptualized, drafted, and edited the manuscript, conducted laboratory analysis, secured funding, and analyzed data. AT analyzed the data, drafted the initial draft together with AJ-G and AG. AJ-G and JC analyzed antibiotic data and contributed to the manuscript. AG coordinated sampling and analyzed microplastics. AdK and AnK conducted metagenomics analysis and edited the manuscript. ÁMÁ coordinated sample collection in Iceland and shipped samples to Norway. MJS performed E. coli SOS analysis, analyzed data, and edited the manuscript. ES and TTT contributed to the project and manuscript editing. ES assisted with sample collection in Finland and sequencing of Finnish samples. AS prepared the wet lab for high-throughput qPCR and managed service procurement in Finland. AdK, ÁMÁ, RL, and TP conceptualized the project, secured funding, supervised the work, and edited the manuscript. All co-authors edited the draft and approved the final version.

### Data availability

All the data produced in the project have been made publicly available in each different publication. All the data included in this paper have been made public as results and supplemental materials.

## Acknowledgements

The authors thank the laboratory personnel at the sites of research for their assistance and coordinating the sample transportation. We also appreciate the contributions of Dr. Kati Räisänen and Odd Gunnar Wikmark throughout the project. Special thanks to the wastewater treatment facility personnel at all surveillance locations for their support in composite sampling and sample transport.

